# In-silico functional prediction of novel tuberculosis pharmacogenetic variants and NAT2 phenotype prediction in African populations

**DOI:** 10.64898/2026.08.17.26360486

**Authors:** Carola Renate Oelofse, Marlo Möller, Caitlin Uren

## Abstract

Tuberculosis (TB) remains a major public health challenge, exerting profound socio-economic burdens and causing debilitating illness in approximately 2.5 million individuals across Africa annually. Optimized large-scale treatment regimens, such as *NAT2*-genotype adjusted dosing, could improve patient outcomes and strengthen healthcare systems. However, fully addressing the complexity of multi-drug TB treatment responses requires consideration of the entire pharmacogenomic (PGx) landscape, particularly within African populations, which are both genetically diverse and critically understudied. In this study, we predict *NAT2* genotypes and phenotypes in specific African populations, and we extend TB PGx research beyond well-established biomarkers. Current bioinformatic prediction tools were used to evaluate individual- and population-specific variation in genotype and next-generation sequencing data from 2,143 individuals across 20 African population groups, spanning ten PGx genes associated with multi-drug TB treatment and response. Most predicted functionally deleterious variants occurred at low frequencies (MAF < 0.01) and were observed in only one of the 20 populations. The Khomani and Nama populations had a distinctly higher proportion of *NAT2* fast metabolizer phenotypes than other African populations, indicating a lower risk of INH overexposure and possibly different dosage requirements in these groups. These findings highlight both the potential and current limitations of functional prediction for absorption, distribution, metabolism and excretion (ADME) variants, and the transferability of their predictive value between African population groups. With the increasing accessibility of next-generation sequencing, alongside the development of comprehensive databases capturing African variation and advances in computational algorithms, the cumulative impact of genetic variation on TB drug response can be more accurately captured, thereby informing precision treatment strategies.

## Introduction

The response to tuberculosis (TB) treatment exhibits substantial variability both across populations and among individuals^1–5^, with toxic drug levels resulting in anti-TB drug-induced liver injury (AT-DILI), amongst other serious adverse reactions (ADRs). Specifically, first-line drugs isoniazid (INH), rifampicin (RIF) and pyrazinamide (PZA) and second-line drug bedaquiline (BDQ) have been implicated in causing ADRs due to accumulation of toxic metabolites. Conversely, sub-therapeutic drug levels can lead to drug resistance, and treatment failure. Genetic variation in absorption, distribution, metabolism, and excretion (ADME) and other pharmacogenomics (PGx) genes account for a significant portion of this complex variability. Importantly, polymorphisms in the *NAT2* gene alter the acetylation rate of INH, leading to predictable rapid or slow metabolism, associated with sub-therapeutic or toxic drug levels, respectively^6^. The benefits and cost effectiveness of *NAT2*-guided INH dosage alterations using allele-specific PCR^7^, or multiplex quantitative PCR assay^8^, on treatment outcomes in clinical application has been demonstrated in clinical settings.

Even so, other association studies investigating TB drug-gene pairs have been less conclusive and no clinical PGx guidelines currently exist for combination TB therapy. The aggregated functional variant frequency, which accounts for the combined directionality and effect sizes of multiple variants, provides a more accurate estimate of expected clinical outcomes than the consideration of individual variants alone^9^, particularly when combination drug therapies increase the number of pharmacogenetic variants that may collectively influence treatment response. For instance, the effect of *CYP2E1* (cytochrome P450 family 2 subfamily E member 1) genotype is evident mostly in *NAT2* slow metabolizers, when alternative pathways of INH metabolism become more predominant^10,11^. In addition, the glutathione S-transferases (GSTM1, GSTP1, GSTT1) enzymes are of increased importance in combination *NAT2/CYP2E1* genotypes^12,13^.

Polymorphisms in other important TB PGx genes alter RIF metabolism, including the solute carrier organic anion transporter family member 1B1 (*SLCO1B1),* arylacetamide deacetylase (*AADAC)* and carboxyl-esterase 2 (*CES2)* genes^14–19^. In addition, cytochrome P450 family 3 subfamily A member 4 (*CYP3A4),* the main enzyme responsible for BDQ metabolism, is strongly induced by RIF treatment, potentially leading to drug-drug interactions^20^. The nuclear pregnane x receptor (PXR) gene *NR1I2* is responsible for regulating the expression of several PGx genes such as *SLCO1B1, CYP3A4* and *CES2*, thereby indirectly influencing the expression and activity of these enzymes, leading to associations with TB treatment outcomes^21^. To date, the ClinPGx database curates more than 200 TB drug-related variant annotations across over 40 genes, linked to broadly different functions such as drug ADME, nuclear hormone receptors, vitamin D pathway, oxidative-stress regulation and immune response.

Notably, most of the ClinPGx assessments of TB drug efficacy and toxicity have historically been conducted primarily in Eurasian cohorts, and knowledge of drug response in African populations - where these treatments are most critically needed – remains limited^22,23^. African genomes encompass the widest spectrum of human genetic diversity, carrying substantially more PGx variants than Europeans^24,25^. Furthermore, determinants of TB drug response differ substantially between populations^22^, with African populations harbouring many population-specific variants, ^19^ and marked differences in allele frequencies, thereby providing important insights into the African PGx landscape^26–28^. Investigations into this diversity have already advanced translational research^29^.

The largest proportion of the variability in drug efficacy and toxicity effects remains unexplained, with approximately 30– 40% of PGx variability stemming from rare variants^9,24^, which by definition are not easily characterized. Furthermore, PGx genes are generally not under strong evolutionary constraint and thus evolutionary conservation alone is a poor predictor of functionality for most bioinformatic tools in ADME genes, despite having substantial effects on protein functionality and drug disposition^9,30,31^. Moderate effects (<10% reduced activity) of some genetic variants are particularly difficult to assess. Structural variation is not only problematic to detect but limited experimental reference data constrains prediction about the functional implications of PGx associated structural variants^31^. The vast multitude of PGx variants makes the experimental approach almost impossibly costly and tedious and thus turning to bioinformatic tools to prioritize variation for further assessment provides a practical and scalable solution.

For this study, we set out to employ in-silico tools to identify novel functionally important TB PGx variation in ten known TB PGx genes (*NAT2, CYP2E1, SLCO1B1, CYP3A4, GSTM1, GSTP1, GSTT1, AADAC, NR1I2 and CES2*) and to characterize functionally important variation in a total of 20 African populations. We determine allele frequency and predict acetylator phenotype information for the key TB pharmacogenomics (PGx) gene, *NAT2*, in the South African Xhosa (XHO), Khomani (KHM), and Nama (NAM) populations. These findings contribute to the currently limited pharmacogenomic data available for these understudied populations and provide further insight into population-specific variation that is influencing TB drug response in a high incidence TB area. Systematic identification, functional annotation, and frequency determination of rare TB PGx variants, supported by bioinformatic and functional studies, will be important for informing PGx testing, optimizing TB drug therapies, and improving clinical outcomes in understudied African populations.

## Methods

Figure 1 summarizes the methodology used in this study to generate a homogenous, high-quality and comprehensive dataset from both WGS and genotyped data sources representing 20 African populations and illustrates the tools and cut-off values applied to further identify and analyse functionally important variation in TB PGx genes.

**Figure 1.**
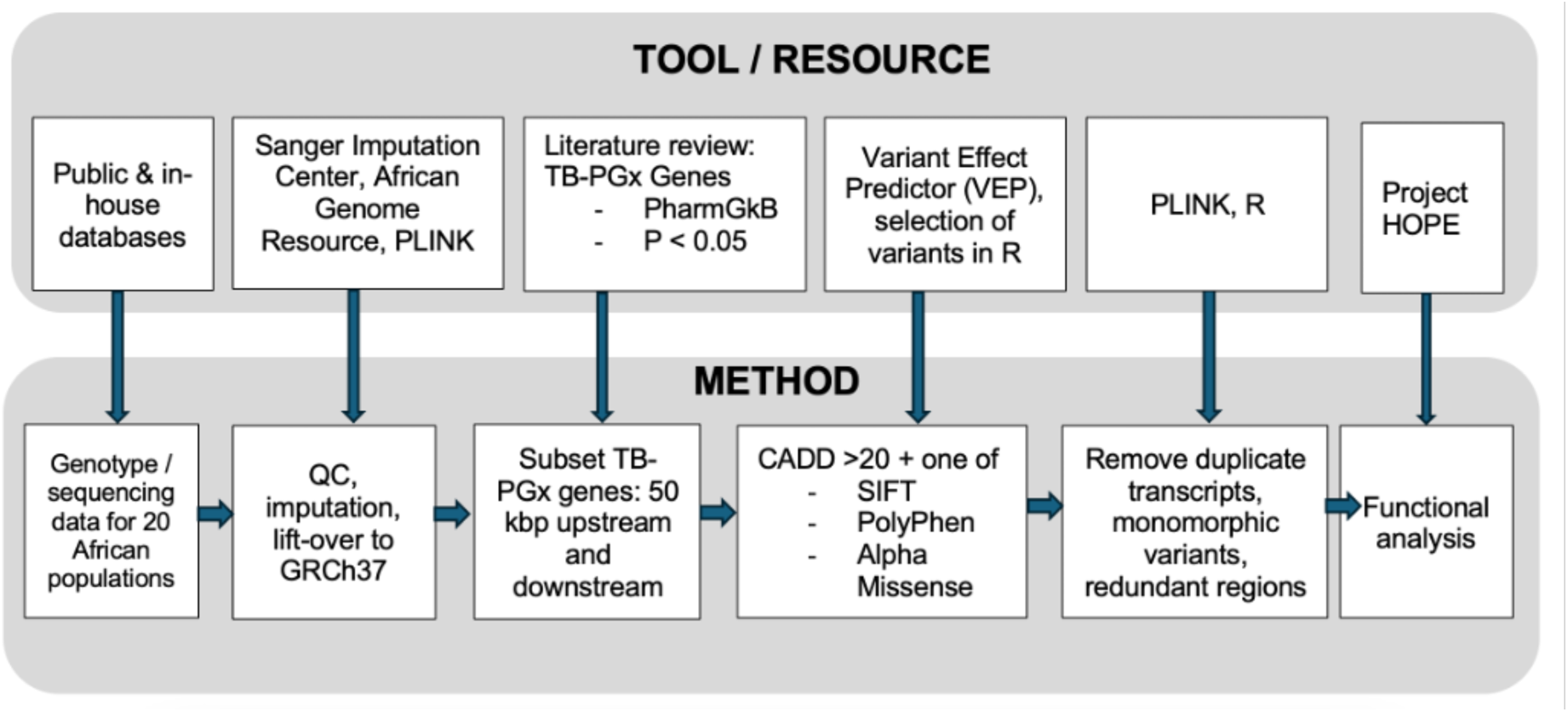
Tools and methodologies used in this study to perform quality control, imputation, TB PGx gene selection, variant effect prediction and in-silico functional analysis in WGS and genotyped datasets in 20 African populations.

### Imputation and Quality Control

Microarray genotyped and whole genome sequenced public and in-house datasets representing 20 African populations across Africa were obtained (Table 1). Briefly, all datasets underwent QC steps using PLINK^32^ (Version 1.9). Indels, variants that did not satisfy Hardy-Weinberg equilibrium, variants for which more than 10% of genotype data was missing, or variants which were monomorphic or tri-allelic, were removed. This was followed by imputation making use of the African Genome Resource (AGR) on the Sanger Imputation platform, as described in a previous publication^29^.

**Table 1.** African populations and dataset sources represented in this study.

| ID | Country | Population / Study Site | Dataset ID | Genotype Technique | Number of Individuals<br>(n = 2143) |
| --- | --- | --- | --- | --- | --- |
| UGA | Uganda | Makerere University | PHS002528.v1.p1<br>(NeuroGAP) | Illumina Global<br>Screening Array | 85 |
| KENK | Kenya | KEMRI Wellcome Trust |  |  | 187 |
| KENM | Kenya | Moi University |  |  | 173 |
| ETH | Ethiopia | Aari, Hamar, Amhara | PHS000449.v2.p1<sup>33</sup> | Human1M-Duo Array | 47 |
| PYG | Cameroon | Baka, Bakola, Bedzan,<br>Lemande, Ngumba, Tikar<br>South | <sup>34</sup> | Illumina 1M SNP Array | 125 |
| NAM | South Africa | Nama | <sup>35</sup> | WGS, Omni Express<br>Plus Array, MEGA Array | 84 |
| XHO |  | Xhosa | Möller/Uren (unpublished<br>data) | WGS | 164 |
| SAC |  | South African Coloured | <sup>36</sup> | H3A Array, Affymetrix<br>500k Array, MEGA<br>Array | 82 |
| KHM |  | Khomani | <sup>37</sup> | Omni Express Plus<br>Array, Omni Express<br>Array, MEGA Array | 185 |
| GHA | Ghana | Ghana | EGAD00010001734<br>(MalariaGEN) | Illumina Omni 2.5M<br>genotyping | 113 |
| MAL | Malawi | Malawi | EGAD00010000903<br>(MalariaGEN) |  | 115 |
| CAM | Cameroon | Cameroon | EGAD00010001740<br>(MalariaGEN) |  | 131 |
| TAN | Tanzania | Tanzania | EGAD00010001743<br>(MalariaGEN) |  | 121 |
| BFA | Burkina<br>Faso | Burkina Faso | EGAD00010001739<br>(MalariaGEN) |  | 122 |
| MZB | Algeria | Mozabite | HGDP<sup>38</sup> | WGS | 27 |
| GWD | Gambia | Gambia Western Division | 1000 GDP<sup>39</sup> | WGS | 104 |
| MSL | Sierra Leone | Mende |  |  | 75 |
| YRI | Nigeria | Yoruba |  |  | 108 |
| LWK | Kenya | Luhya |  |  | 82 |
| EGY | Egypt | Egyptian | SGDP <sup>40</sup> | Affymetrix Human Origins Array | 13 |

The AGR was shown to provide the highest imputation accuracy in Southern African populations, filling in missing genotypes even in low-frequency variation^29^. Adhering to an INFO score threshold of 0.6, genotype imputation quality score filtering was applied to ensure consistent data quality, completeness and reliability across datasets derived from diverse sources and genotyping or sequencing platforms (Table 1).

### Variant Prioritization

For this study, ten TB PGx genes were prioritized based on evidence curated from published literature available through the ClinPGx resource (Table 2). Gene selection was guided by the presence of repeated statistically significant associations (p < 0.05) across multiple population groups, linking genetic variants to variability in anti-TB drug pharmacokinetics and clinically relevant treatment outcomes, including AT-DILI. Gene regions of interest, including 50 kbp upstream and downstream, were extracted using PLINK^32^. Resulting variant call format (VCF) files were uploaded to the Ensembl Variant Effect Predictor (VEP) tool^41^, which provides user-friendly access to top performing functional analysis tools CADD (Combined Annotation Dependent Depletion)^42^, PolyPhen (Polymorphism Phenotyping v2)^43^, SIFT (Sorting Intolerant From Tolerant)^44^ and Alpha Missense^45^. These tools have specifically been assessed for their ability to predict function in pharmacogenes, with Alpha Missense producing the most specific structural predictions^46^. Project HOPE incorporates sequence annotations from UniProt and 3D protein structural information to predict the effect of point mutations on protein domains, active sites and chemical interaction with substrates, using a variety of databases and online resources to produce 3D figures and animations^47^. This tool was chosen to gain a more comprehensive insight into the structural effect these variants could specifically have on the catalytic function of enzymes.

**Table 2.** Genes associated with TB drug treatment outcome and/or AT-DILI, not exhaustively representing all the variations in these genes previously associated with TB PGx in current literature. Genes (n=10) were prioritized for in-silico functional analysis.

| Gene | SNPs in target gene associated with TB PGx / AT-DILI | Significance | Reference | Drug |
| --- | --- | --- | --- | --- |
| AADAC | rs1803155 (841G>A) | P = 0.0015 | PMID: 37732197<sup>48</sup> | RIF |
| CES2 | CES2 c.-22263A>G g.738A>G |  | PMID: 35693129<sup>49</sup> | RIF |
| CYP2E1 | CYP2E1 (rs2031920) | OR: 0.16; P = 0.049 | PMID: 35751408<sup>13</sup> | INH |
|  | (rs2031920) | OR: 1.18 (95% CI: 0.82–1.71) | PMID: 34154362<sup>50</sup> |  |
| CYP3A4 | *5 (rs55901263) | P < 0.05 | PMID: 34872459<sup>20</sup> | RIF |
| GSTM1 | null genotype | OR: 1.169 (95% CI: 1.028–1.330); P = 0.038 | PMID: 31378997<sup>51</sup> | INH |
| GSTP1 | rs1695 | OR: 2.10 (95%CI: 1.17–3.76); P = 0.012 | PMID: 27281183<sup>52</sup> | INH |
| GSTT1 | null genotype | OR: 3.44 (95%CI: 1.01 to 11.71); P = 0.048 | PMID: 33981901<sup>12</sup> | INH |
| **NAT2** | NAT2 acetylator phenotypes |  | PMID: 37454203<sup>53</sup> | INH |
|  | NAT2 acetylator phenotypes | OR: 5.97 (95% CI: 1.38–25.76); *P* = 0.02 | PMID: 39727196<sup>54</sup> | INH |
|  | NAT2 acetylator phenotypes | SHR = 3.05; (95% CI: 1.17–7.95); *P* = 0.02 | PMID: 40332508<sup>55</sup> | INH |
|  | NAT2*5 and NAT2*6 | OR: 4.2 (95%CI: 1.1–15.2); *P* = 0.017 | PMID: 39085767<sup>56</sup> | INH |
|  | NAT2*4, NAT2*5A, NAT2*6A, and NAT2*13A |  | PMID: 28959610<sup>57</sup> | INH |
|  | NAT2 acetylator phenotypes |  | PMID: 38629592<sup>5</sup> | INH |
| **NR1I2 (PXR)** | rs7643645 |  | PMID: 30778091<sup>21</sup> | RIF |
| **SLCO1B1** | *15 (rs4149056) | OR: 1.74 (95% CI: 1.04–2.90); *P* = 0.034 | PMID: 25498879<sup>15</sup> | RIF |
|  | haplotypes |  | PMID: 28900877<sup>58</sup> |  |
| **TOTAL** |  |  |  |  |

### Functional Variant Effect Prediction

Files predicting functional effects of SNPs were merged in R (version 4.4.1), and the following previously used^59^ filtering cut-offs were applied: CADD scores > 20 and either one of the three other criteria, PolyPhen “probably damaging” (> 0.85), SIFT “deleterious” (< 0.05) and Alpha Missense (>0.564). CADD compares observed human variants with those under investigation, assigning a score based on conservation, protein impact and transcript-level effects. A CADD PHRED score of >20 means that a variant is 99% more deleterious than all possible variants and was thus used as the threshold in our study. Combining the CADD score with either SIFT, PolyPhen or Alpha Missense resulted in the selection of highly likely functionally important variants that would have an effect based either on conservation principles (SIFT), sequence homology, protein 3D structure (including active binding sites) (PolyPhen) and deep learning (Alpha- Missense). MANE (matched annotation from NCBI and EMBL-EBI) transcripts were selected, and no frequency filters were applied. Allele frequencies for functionally important variants were called for all populations using PLINK^32^. For comparison of African allele frequencies with other global populations, allele frequencies for South Asian (SAS) and European and Non-Finnish European (EUR/NFE) populations were obtained from gnomAD (v4.1.0).

### NAT2 allele calling

We assessed star allele haplotype frequencies for *NAT2* using PyPGx (version 0.25.0), which identifies alleles by phasing and comparing combinations of allele-defining variants against a database^36^. PyPGx is compatible with both genotyped and sequenced data. Recent, fundamental changes to the legacy *NAT2* nomenclature database since March 2024^60^ have not yet been accounted for by PyPGx v.0.25.0 or other allele and phenotype callers such PGxPOP v.0.1.0^61^, or Illumina Dragen Software v.4.4. Although StellarPGx^62^ and ALDY^63^ are updated to current nomenclature, these allele callers are only suitable for sequenced data and are therefore not compatible with the datasets used in this study. As described by the latest Clinical Pharmacogenetics Implementation Consortium (CPIC) consensus-based framework, “star alleles” interchangeably refer to either a single allele-defining genetic variant, or a defined combination of variants (haplotype)^64^. Thus, for completeness and greater comprehensiveness, we focus on the predicted function of the allele-defining variant, as well as both current (https://www.pharmvar.org/gene/NAT2) and legacy (https://nat.mbg.duth.gr/Human_NAT2_alleles.htm) star allele nomenclature^60^. Diplotypes were called using PyPGx and predicted acetylator phenotypes assigned as follows: rapid metabolizer (two functional alleles), intermediate metabolizer (one functional and one decreased-function allele) and poor (slow) metabolizer (two non-functional alleles) or indeterminate (one indeterminate or unknown-function allele) as per the current genotype-phenotype CPIC guidelines (https://www.clinpgx.org/page/nat2RefMaterials, accessed on 30 May 2026).

## Results

### VEP Results and Variant Filtering

Across the 20 African populations, an average of ∼11 873 variants were identified in *NAT2*, *CYP2E1*, *SLCO1B1, AADAC, CYP3A4, GSTM1, GSTP1, GSTT1*, *NR1I2* and *CES2* combined. The percentage of variants without previously assigned rs IDs varied between the populations, with the lowest number in Esan, Nigeria (ESN) (2.9%) and the highest number of unassigned variants in the NAM (13.2%). Across all populations, the majority of variants were intronic (average 61%), while most of coding consequences were categorized as missense variants (average 60%). After *in-silico* analysis, around 1181 variants remained in the combined dataset. After removing monomorphic sites and duplications across all populations, 34 variants remained in the dataset and were divided according to their presence (Table 3) or absence (Table 4) in PubMed records.

**Table 3.**
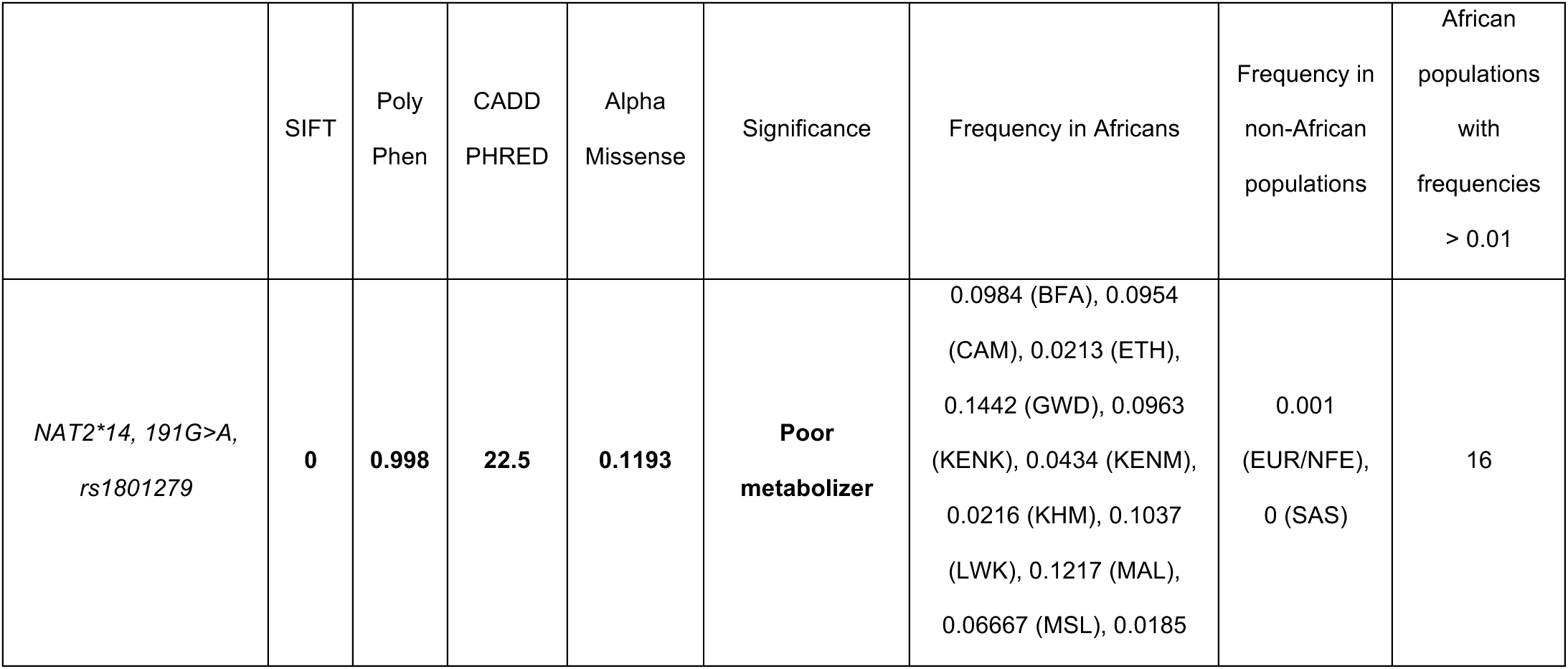
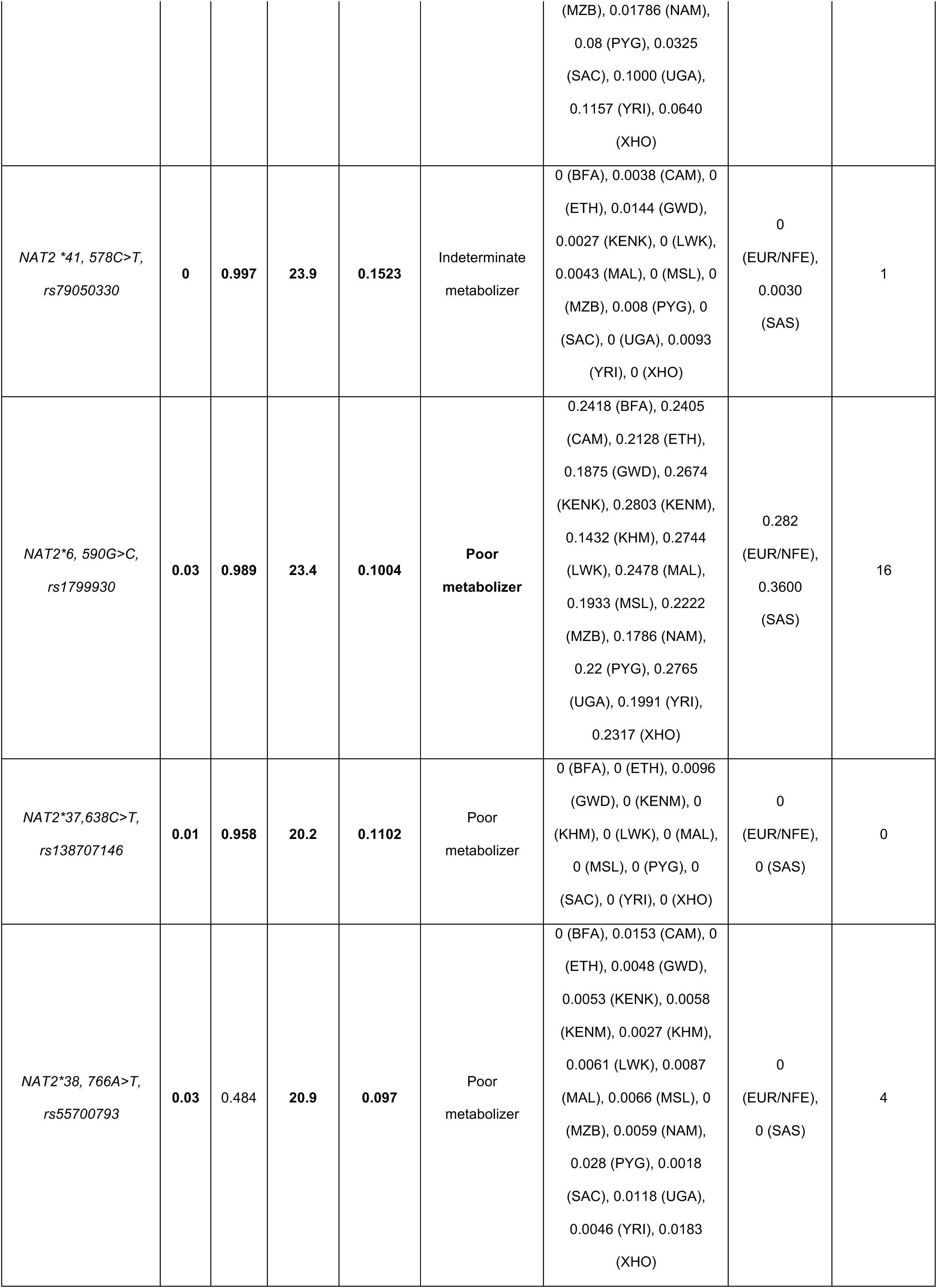
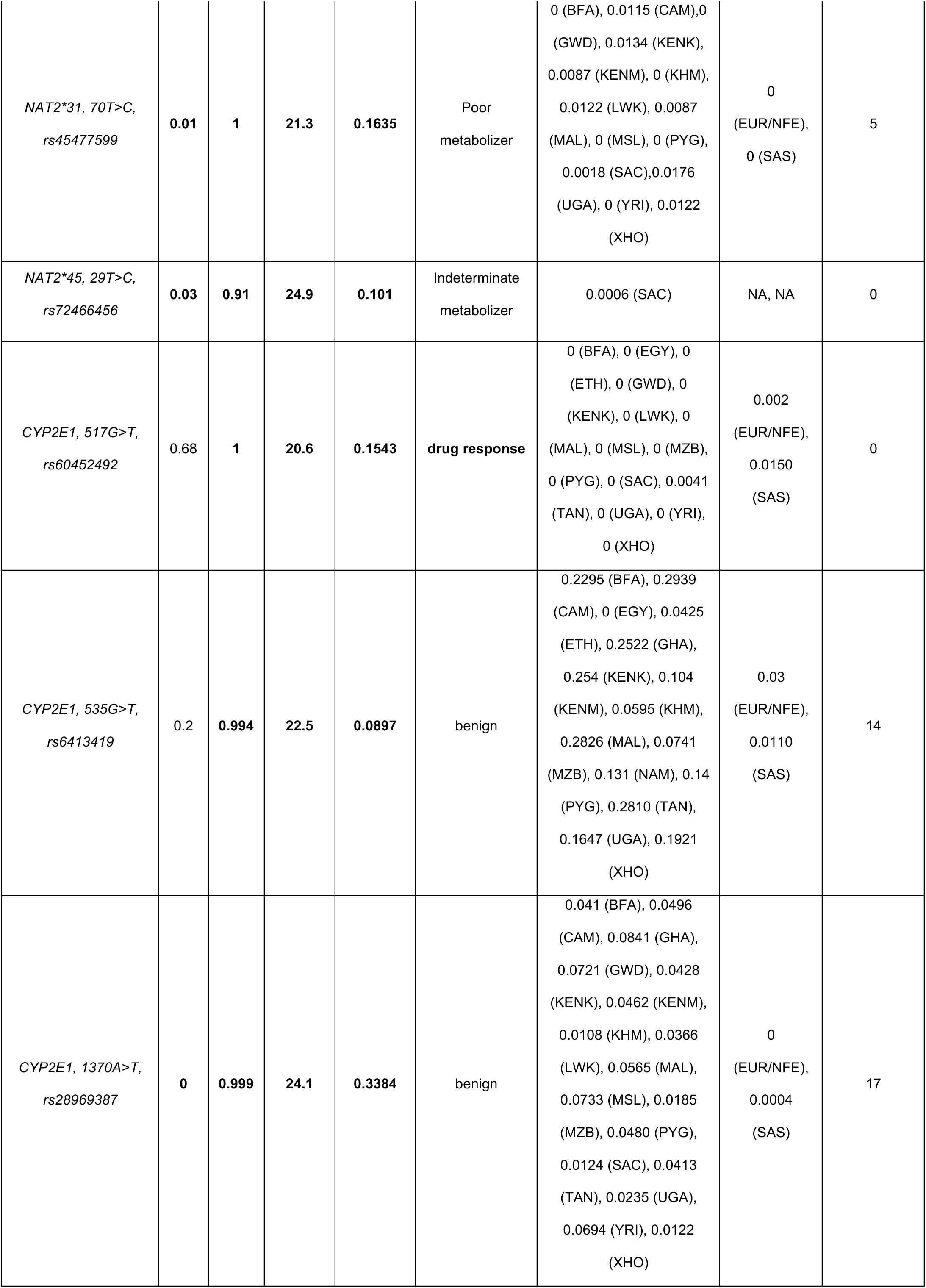
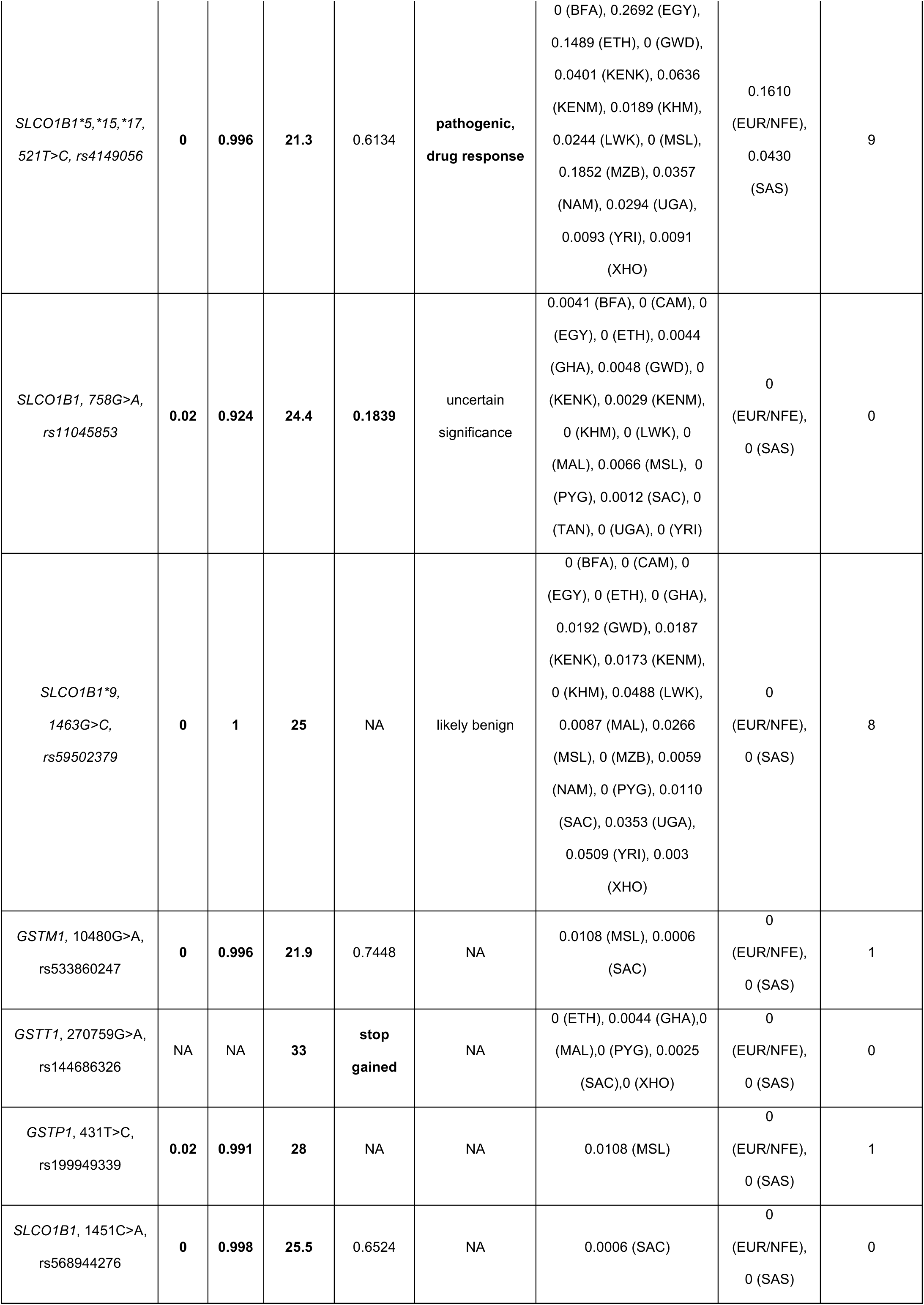
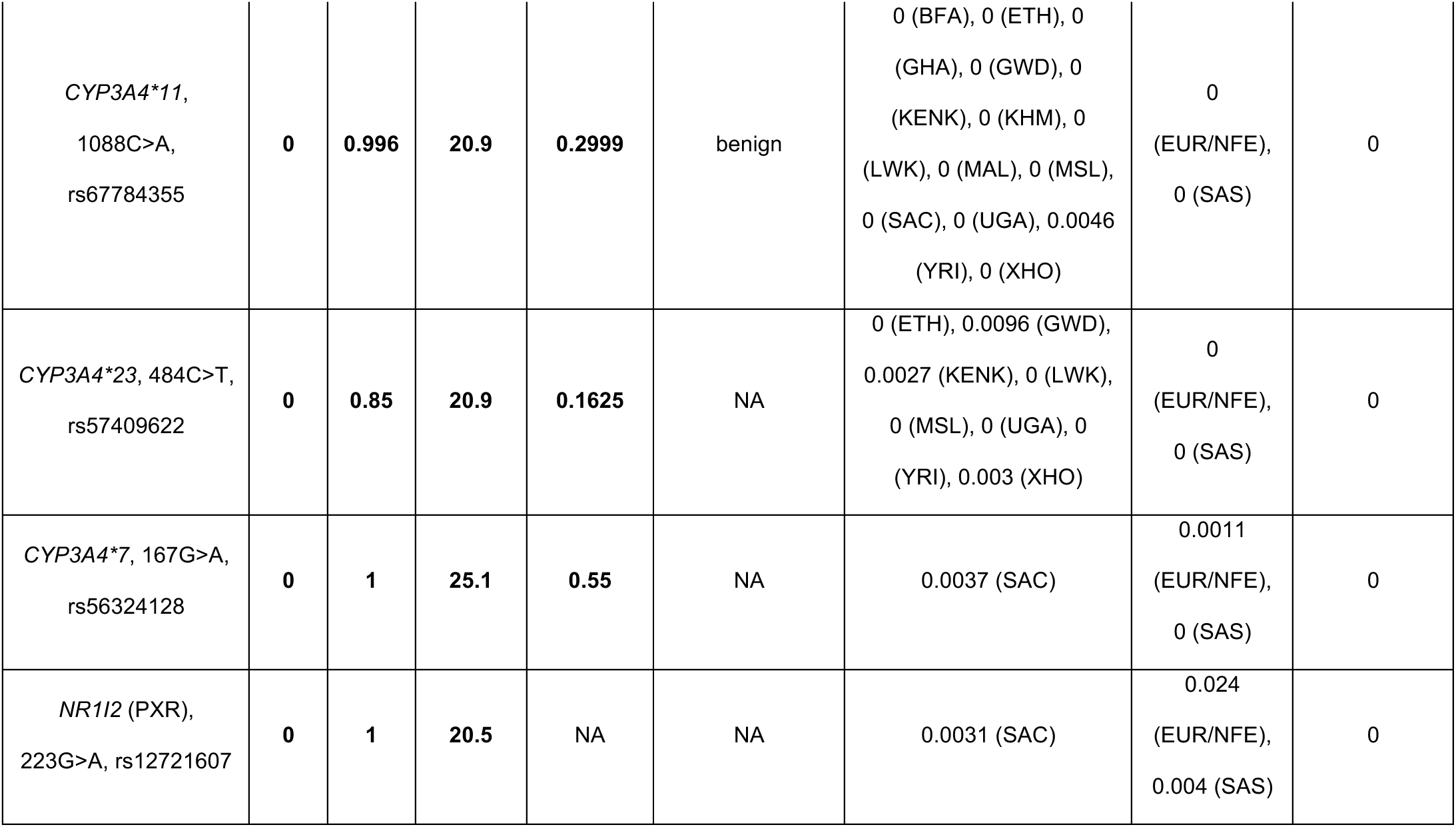
Variants with PubMED records, in prioritized TB PGx genes predicted to have a functional effect. SIFT < 0.05 = deleterious. PolyPhen > 0.85 = damaging. Alpha Missense > 0.564 = likely pathogenic. CADD > 20 = damaging. In-silico predictive values reaching these cut-offs are shown in bold.

**Table 4.** Variants without PubMED records, in prioritized TB PGx genes predicted to have a functional effect. SIFT < 0.05 = deleterious. PolyPhen > 0.85 = damaging. Alpha Missense > 0.564 = likely pathogenic. CADD > 20 = damaging. In-silico predictive values reaching these cut-offs are shown in bold.

|  | SIFT | PolyPhen | CADD<br>PHRED | Alpha<br>Missense | Frequency in African<br>populations | Frequency in non-African<br>populations | African Populations<br>with frequencies ><br>0.01 |
| --- | --- | --- | --- | --- | --- | --- | --- |
| SLCO1B1, 242T>C,<br>rs1353535924 | **0** | **0.996** | **25.7** | 0.4321 | 0.0185 (MZB) | 0 (NFE/EUR), 0 (SAS) | 1 |
| CYP2E1, 646C>G,<br>rs1194608147 | 0.05 | 0.767 | **23.4** | 0.2624 | 0 (BFA), 0 (KENK), 0.0029 (KENM), 0 (KHM), 0 (MAL), 0 (PYG), 0 (UGA), 0 (XHO) | 0 (NFE/EUR), 0 (SAS) | 0 |
| CYP3A4, 1072A>G,<br>rs754968125 | **0** | 0.822 | **24.3** | 0.3772 | 0 (GHA), 0.0027 (KENK), 0 (KENM), 0 (XHO) | 0 (NFE/EUR), 0 (SAS) | 0 |
| NR1I2 (PXR),<br>647G>A,<br>rs768816150 | **0.01** | **1** | **26.1** | NA | 0.0053 (KENM), 0 (MZB), 0 (XHO) | 0 (NFE/EUR), 0.0002 (SAS) | 0 |
| NR1I2 (PXR),<br>700G>A,<br>rs144833620 | **0.01** | **1** | **31** | NA | 0.0029 (KENM), 0 (MZB), 0.0012 (SAC), 0 (XHO) | 0 (NFE/EUR), 0.0001 (SAS) | 0 |
| NR1I2 (PXR),<br>720G>T,<br>rs371067095 | **0** | **1** | **22.9** | NA | 0.0185 (MZB) | 0.0001 (NFE/EUR), 0 (SAS) | 1 |
| NR1I2 (PXR),<br>1099G>C,<br>rs1194240746 | 0.02 | 0.309 | 23.7 | NA | 0.0027 (KENK) | 0 (NFE/EUR), 0<br>(SAS) | 0 |
| AADAC, 271G>C,<br>rs1716042355 | 0 | 0.977 | 24 | 0.5316 | 0.0041 (BFA), 0 (GHA),<br>0.0029 (KENM), 0 (KHM), 0<br>(MAL), 0 (TAN), 0 (XHO) | 0 (NFE/EUR), 0<br>(SAS) | 0 |
| AADAC, 274C>T,<br>rs760383318 | 0 | 0.996 | 27.1 | 0.2645 | 0 (BFA), 0 (GHA), 0 (KENM),<br>0.0135 (KHM), 0 (MAL), 0<br>(PYG), 0 (TAN), 0 (XHO) | 0 (NFE/EUR), 0<br>(SAS) | 1 |
| CES2, 467A>G,<br>rs1048849276 | 0.02 | 0.204 | 22.8 | NA | 0 (GHA), 0 (KENM), 0<br>(MSL), 0.0043 (MZB) | 0 (NFE/EUR), 0<br>(SAS) | 0 |
| CES2, 621G>A,<br>rs565364625 | 0.02 | 0.998 | 27.7 | NA | 0.0006 (NAM) | 0 (NFE/EUR), 0<br>(SAS) | 0 |
| CES2, 725G>A,<br>rs112732183 | 0.01 | 0.978 | 23.4 | NA | 0 (GHA), 0 (KENM), 0.1216<br>(LWK), 0.0488 (MSL)<br>, 0.0043 (MZB), 0.1071<br>(XHO) | 0 (NFE/EUR), 0<br>(SAS) | 3 |
| CES2, 662G>A,<br>rs537562880 | 0 | 0.998 | 24.8 | NA | 0.0185 (MZB) | 0 (NFE/EUR), 0<br>(SAS) | 1 |

For 34 variants across 20 populations, (a total of 680 calls), frequency data was not available (NA) for most calls (n=369), indicating that over half of the variants that were typed in one population were not typed in most others (Figure 2B). In datasets sourced from 1000 Genomes (Luhya Western Kenya (LWK), Gambia Western Division (GWD), Yoruba Nigeria (YRI), ESN, Ghana (GHA) and Mende Sierra Leone (MSL)), the slow *NAT2*5* allele did not meet quality control criteria. Moreover, low sample numbers in Mozabite (MZB) (n=27), Ethiopian (ETH) (n=47) and Egyptian (EGY) (n=13) datasets constrained reliable allele prediction. These populations were thus excluded from *NAT2* phenotype prediction (Figure 3B, Supplementary Table 2).

**Figure 2.**
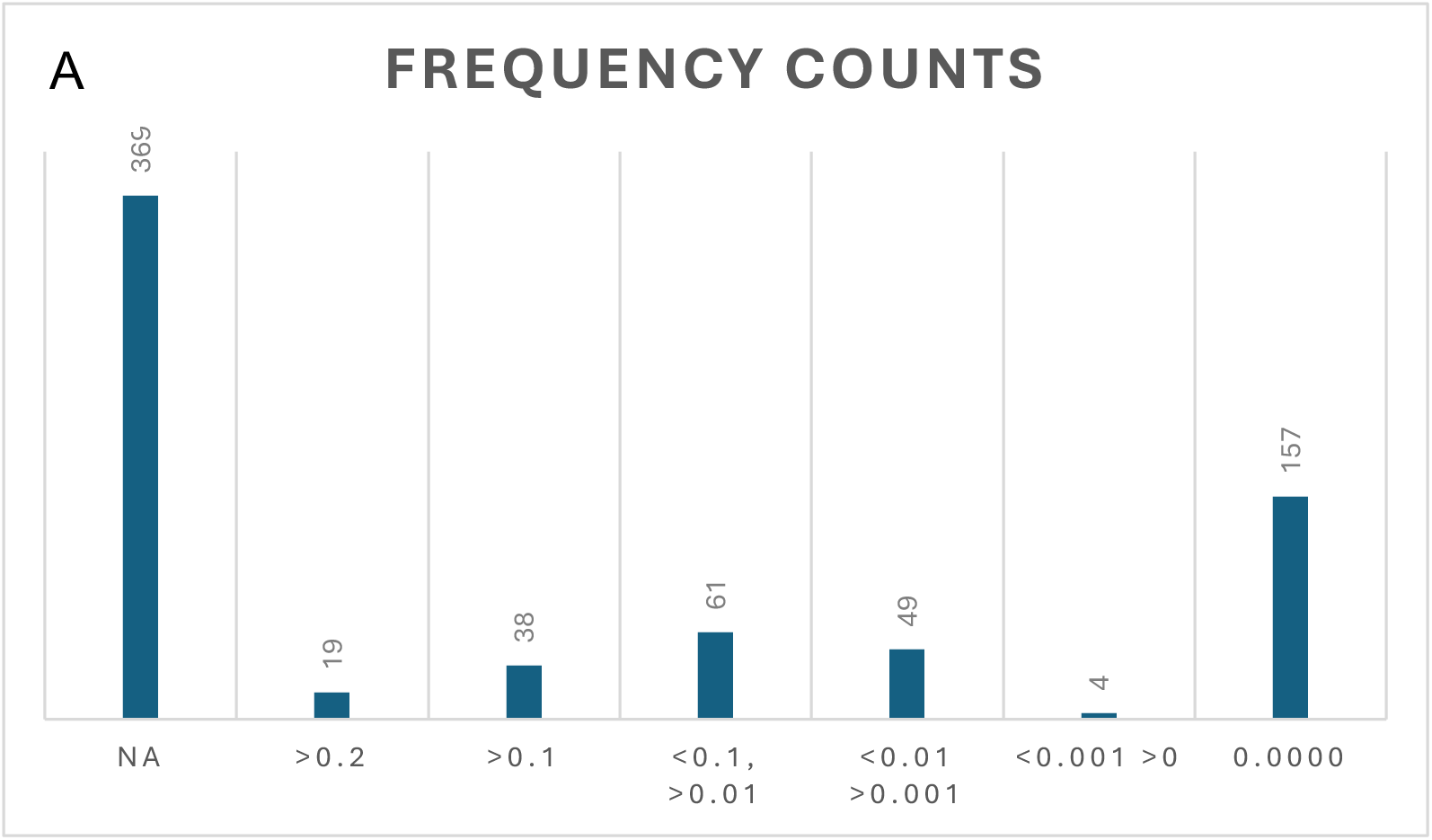
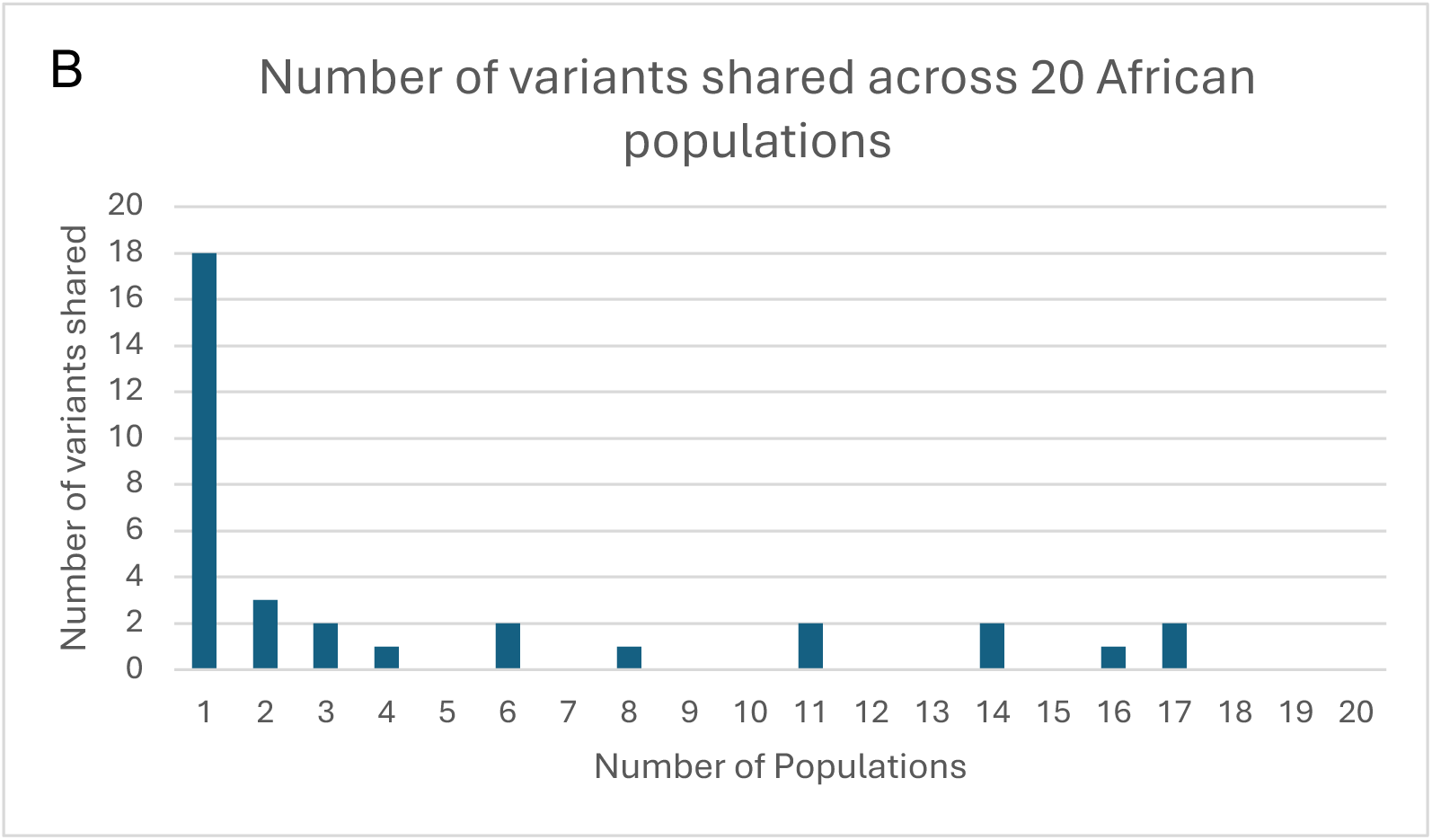
A) Allele frequency distribution binned across 20 African populations in 34 functionally important variants. For the majority of calls, no frequency information was available (NA). B) Number of instances functionally important variants (n=34) are shared across 20 African populations. No single variant was shared by more than 17 populations.

**Figure 3.**
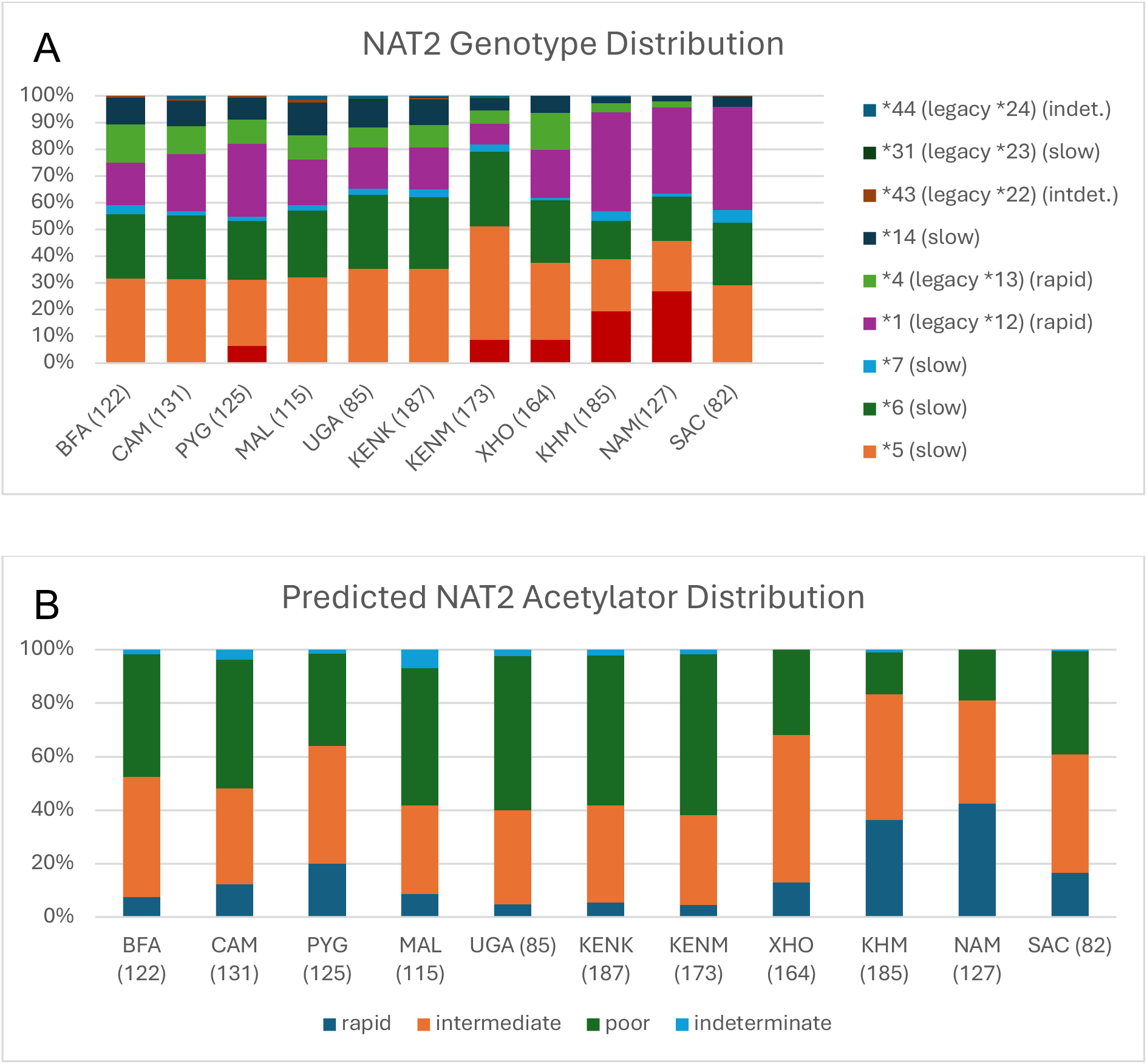
A) NAT2 genotypes across African populations. Populations LWK, YRI, GWD, TAN and MSL were excluded because the allele-defining variant NAT2*5 (rs1801280) did not meet the INFO-score threshold of > 0.6 in these populations. EGY, MZB and ETH were excluded because of small sample numbers. B) NAT2 acetylator distribution, predicted using PyPGx and the CPIC NAT2 genotype-phenotype table (https://www.clinpgx.org/page/nat2RefMaterials). BFA=Burkina Faso, CAM=Cameroon, ETH=Ethiopia, GWD=Gambia Western Division, KENK=Kenya KEMRI, KENM=Kenya Moi, KHM = Khomani, LWK=Luhya Western Kenya, MAL=Malawi, MSL=Mende Sierra Leone, MZB=Mozabite, PYG=Pygmy, SAC=South African Coloured, UGA=Uganda, XHO=Xhosa.

Of the variant calls that were typed (n=311) the majority (n=157) had a frequency of zero calls, 61 calls had a frequency between < 1% and >0.1%, and 49 calls fell into a frequency category between < 0.1% and > 0.01% (Figure 2A). A total of 18 of the variant entries across all 20 populations were more frequent than 0.2% (Figure 2A). Approximately half of all identified variants occurred in only one of the 20 populations, with none of the variants occurring in all populations (Figure 2B). Most variants had a CADD PHRED score >20 and <25, while only two variants exceeded a score >30 (Tables 3 and 4).

### NAT2

The KHM and NAM have significantly higher proportions of the fast acetylator *NAT2*4.001* (legacy NAT2*4) allele (Figure 3A, Supplementary Table 1). Conversely, the non-functional *NAT2*14* allele occurs in all 20 African populations investigated here (Table 5), but at low proportions in the NAM (1.93), KHM (2.16) and XHO (6.42) (Figure 3A, Supplementary Table 1). This combination of alleles contributes to a comparatively higher number of fast acetylator phenotypes in these South African populations (Figure 3B).

**Table 5.**
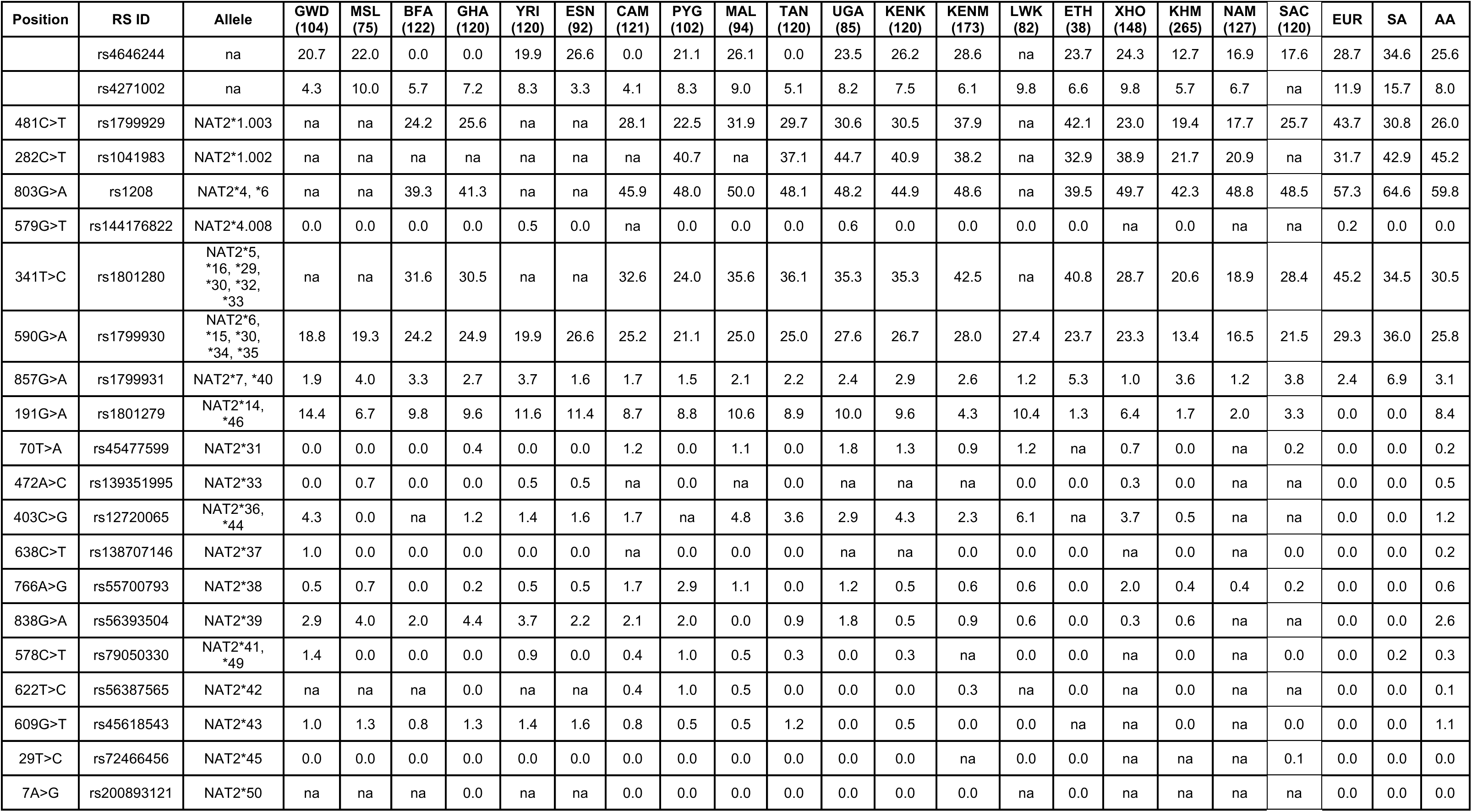
Frequencies of current NAT2 allele-defining SNPs across African populations in this study. Populations are ordered geographically from Western-Eastern-Southern Africa. GWD=Gambia Western Division, MSL=Mende Sierra Leone, BFA=Burkina Faso, GHA=Ghana, YRI=Yoruba Nigeria, ESN=Esan Nigeria, CAM=Cameroon, PYG=Pygmy, MAL=Malawi, TAN=Tanzania, UGA=Uganda, KENK=Kenya Kemri, KENM=Kenya Moi, LWK=Luhya Western Kenya, ETH=Ethiopia, XHO=Xhosa, KHM=Khomani, NAM=Nama, SAC=South African Coloured. Frequency information for European (EUR), SA (South Asian) and AA (African American) populations was sourced from the gnomAD (v4.1.0) resource.

| Position | RS ID | Allele | GWD<br>(104) | MSL<br>(75) | BFA<br>(122) | GHA<br>(120) | YRI<br>(120) | ESN<br>(92) | CAM<br>(121) | PYG<br>(102) | MAL<br>(94) | TAN<br>(120) | UGA<br>(85) | KENK<br>(120) | KENM<br>(173) | LWK<br>(82) | ETH<br>(38) | XHO<br>(148) | KHM<br>(265) | NAM<br>(127) | SAC<br>(120) | EUR | SA | AA |
| --- | --- | --- | --- | --- | --- | --- | --- | --- | --- | --- | --- | --- | --- | --- | --- | --- | --- | --- | --- | --- | --- | --- | --- | --- |
|  | rs4646244 | na | 20.7 | 22.0 | 0.0 | 0.0 | 19.9 | 26.6 | 0.0 | 21.1 | 26.1 | 0.0 | 23.5 | 26.2 | 28.6 | na | 23.7 | 24.3 | 12.7 | 16.9 | 17.6 | 28.7 | 34.6 | 25.6 |
|  | rs4271002 | na | 4.3 | 10.0 | 5.7 | 7.2 | 8.3 | 3.3 | 4.1 | 8.3 | 9.0 | 5.1 | 8.2 | 7.5 | 6.1 | 9.8 | 6.6 | 9.8 | 5.7 | 6.7 | na | 11.9 | 15.7 | 8.0 |
| 481C>T | rs1799929 | NAT2*1.003 | na | na | 24.2 | 25.6 | na | na | 28.1 | 22.5 | 31.9 | 29.7 | 30.6 | 30.5 | 37.9 | na | 42.1 | 23.0 | 19.4 | 17.7 | 25.7 | 43.7 | 30.8 | 26.0 |
| 282C>T | rs1041983 | NAT2*1.002 | na | na | na | na | na | na | na | 40.7 | na | 37.1 | 44.7 | 40.9 | 38.2 | na | 32.9 | 38.9 | 21.7 | 20.9 | na | 31.7 | 42.9 | 45.2 |
| 803G>A | rs1208 | NAT2*4, *6 | na | na | 39.3 | 41.3 | na | na | 45.9 | 48.0 | 50.0 | 48.1 | 48.2 | 44.9 | 48.6 | na | 39.5 | 49.7 | 42.3 | 48.8 | 48.5 | 57.3 | 64.6 | 59.8 |
| 579G>T | rs144176822 | NAT2*4.008 | 0.0 | 0.0 | 0.0 | 0.0 | 0.5 | 0.0 | na | 0.0 | 0.0 | 0.0 | 0.6 | 0.0 | 0.0 | 0.0 | 0.0 | na | 0.0 | na | na | 0.2 | 0.0 | 0.0 |
| 341T>C | rs1801280 | NAT2*5,<br>*16, *29,<br>*30, *32,<br>*33 | na | na | 31.6 | 30.5 | na | na | 32.6 | 24.0 | 35.6 | 36.1 | 35.3 | 35.3 | 42.5 | na | 40.8 | 28.7 | 20.6 | 18.9 | 28.4 | 45.2 | 34.5 | 30.5 |
| 590G>A | rs1799930 | NAT2*6,<br>*15, *30,<br>*34, *35 | 18.8 | 19.3 | 24.2 | 24.9 | 19.9 | 26.6 | 25.2 | 21.1 | 25.0 | 25.0 | 27.6 | 26.7 | 28.0 | 27.4 | 23.7 | 23.3 | 13.4 | 16.5 | 21.5 | 29.3 | 36.0 | 25.8 |
| 857G>A | rs1799931 | NAT2*7, *40 | 1.9 | 4.0 | 3.3 | 2.7 | 3.7 | 1.6 | 1.7 | 1.5 | 2.1 | 2.2 | 2.4 | 2.9 | 2.6 | 1.2 | 5.3 | 1.0 | 3.6 | 1.2 | 3.8 | 2.4 | 6.9 | 3.1 |
| 191G>A | rs1801279 | NAT2*14,<br>*46 | 14.4 | 6.7 | 9.8 | 9.6 | 11.6 | 11.4 | 8.7 | 8.8 | 10.6 | 8.9 | 10.0 | 9.6 | 4.3 | 10.4 | 1.3 | 6.4 | 1.7 | 2.0 | 3.3 | 0.0 | 0.0 | 8.4 |
| 70T>A | rs45477599 | NAT2*31 | 0.0 | 0.0 | 0.0 | 0.4 | 0.0 | 0.0 | 1.2 | 0.0 | 1.1 | 0.0 | 1.8 | 1.3 | 0.9 | 1.2 | na | 0.7 | 0.0 | na | 0.2 | 0.0 | 0.0 | 0.2 |
| 472A>C | rs139351995 | NAT2*33 | 0.0 | 0.7 | 0.0 | 0.0 | 0.5 | 0.5 | na | 0.0 | na | 0.0 | na | na | na | 0.0 | 0.0 | 0.3 | 0.0 | na | na | 0.0 | 0.0 | 0.5 |
| 403C>G | rs12720065 | NAT2*36,<br>*44 | 4.3 | 0.0 | na | 1.2 | 1.4 | 1.6 | 1.7 | na | 4.8 | 3.6 | 2.9 | 4.3 | 2.3 | 6.1 | na | 3.7 | 0.5 | na | na | 0.0 | 0.0 | 1.2 |
| 638C>T | rs138707146 | NAT2*37 | 1.0 | 0.0 | 0.0 | 0.0 | 0.0 | 0.0 | na | 0.0 | 0.0 | 0.0 | na | na | 0.0 | 0.0 | 0.0 | na | 0.0 | na | 0.0 | 0.0 | 0.0 | 0.2 |
| 766A>G | rs55700793 | NAT2*38 | 0.5 | 0.7 | 0.0 | 0.2 | 0.5 | 0.5 | 1.7 | 2.9 | 1.1 | 0.0 | 1.2 | 0.5 | 0.6 | 0.6 | 0.0 | 2.0 | 0.4 | 0.4 | 0.2 | 0.0 | 0.0 | 0.6 |
| 838G>A | rs56393504 | NAT2*39 | 2.9 | 4.0 | 2.0 | 4.4 | 3.7 | 2.2 | 2.1 | 2.0 | 0.0 | 0.9 | 1.8 | 0.5 | 0.9 | 0.6 | 0.0 | 0.3 | 0.6 | na | na | 0.0 | 0.0 | 2.6 |
| 578C>T | rs79050330 | NAT2*41,<br>*49 | 1.4 | 0.0 | 0.0 | 0.0 | 0.9 | 0.0 | 0.4 | 1.0 | 0.5 | 0.3 | 0.0 | 0.3 | na | 0.0 | 0.0 | na | 0.0 | na | 0.0 | 0.0 | 0.2 | 0.3 |
| 622T>C | rs56387565 | NAT2*42 | na | na | na | 0.0 | na | na | 0.4 | 1.0 | 0.5 | 0.0 | 0.0 | 0.0 | 0.3 | na | 0.0 | na | 0.0 | na | na | 0.0 | 0.0 | 0.1 |
| 609G>T | rs45618543 | NAT2*43 | 1.0 | 1.3 | 0.8 | 1.3 | 1.4 | 1.6 | 0.8 | 0.5 | 0.5 | 1.2 | 0.0 | 0.5 | 0.0 | 0.0 | na | na | 0.0 | na | 0.0 | 0.0 | 0.0 | 1.1 |
| 29T>C | rs72466456 | NAT2*45 | 0.0 | 0.0 | 0.0 | 0.0 | 0.0 | 0.0 | 0.0 | 0.0 | 0.0 | 0.0 | 0.0 | 0.0 | na | 0.0 | 0.0 | na | na | na | 0.1 | 0.0 | 0.0 | 0.0 |
| 7A>G | rs200893121 | NAT2*50 | na | na | na | 0.0 | na | na | 0.0 | 0.0 | 0.0 | 0.0 | 0.0 | 0.0 | 0.0 | na | 0.0 | na | na | na | na | 0.0 | 0.0 | 0.0 |

Three slow acetylator alleles, *NAT2*31 (*70T>C, rs45477599, legacy *NAT2*23*), *NAT2*37* (638C>T) and *NAT2*38* (766A>T), have been identified here as functionally relevant and occur at frequencies of >1% across several African populations in this study (Table 5), but do not occur at significant frequencies in EUR/NFE and SAS populations (Table 5). Three alleles of indeterminate acetylation phenotype were identified in several African populations. Both the *NAT2*43* (609G>T, rs55700793, legacy *NAT2*22*) and *NAT2*44* (403C>G, rs12720065, legacy *NAT2*24*) were not found functionally relevant in our study. *NAT2*36* also carries the allele-defining 403C>G variant in combination with other variants but could not be differentiated using PyPGx. The indeterminate function *NAT2*45* allele was distinguished in only the SAC population in this study and was predicted to have a functional effect in-silico (Table 3).

### SLCO1B1

Three uncharacterized variants, which are more frequent in African populations than non-African populations, were noteworthy, according to our functional analysis. Firstly, the *SLCO1B1* variant 242T>C (rs1353535924) could be of interest in the MZB population (Table 3). Project HOPE indicated this variant to be smaller, less hydrophobic, and likely pathogenic (Supplementary Table 3). Secondly, the *SLCO1B1* SNP 1451C>A (rs568944276) was predicted to bear a functional effect (Table 3) but tolerated experimentally^65^. Thirdly, the *SLCO1B1* variant 758G>A (rs11045853) occurs in six of the African populations listed, but at low frequencies, the highest in the MSL population (0.006). According to Project HOPE, this variant does not cause loss of function to the protein but might interfere with its ability to interact with other molecules (Supplementary Table 3).

### CYP2E1

Although *CYP2E1* 535G>T (rs6413419) is predicted to be deleterious and probably damaging by SIFT and PolyPhen (Table 3), current literature describes it as functionally normal^66^. The *CYP2E1* variant 517G>T occurs only in one of the 20 populations and is thought to be functionally normal or to increase hydroxylation activity^66^. The *CYP2E1* 1370A>T (rs28969387) variant occurs in 17 populations at low frequencies, is practically absent in EUR/NFE and SAS populations and is predicted to have a functional impact *in silico* (Supplementary Table 3). The *CYP2E1* 646 C>G (rs1194608147) variant was detected exclusively in the KENM (0.0029) and Project HOPE analysis suggests it exerts a functional effect (Supplementary Table 3).

### GST

The GSTT1 variant 270759G>A (rs144686326) results in a stop gained, but its frequency in most populations is very low (Table 3). Similarly, the *GSTM1* variant 10480G>A (rs533860247) was predicted to destabilize protein function (Table 3, Supplementary Table 3). *GSTP1* 431T>C (rs199949339) is predicted to be less pathogenic (Table 3, Supplementary Table 3) but was only identified in the KHM in this study (0.01087). All three of these variants do not occur at significant frequencies in the EUR/NFE and SAS populations, according to gnomAD (v4.1.0).

### Additional genes

Several variants with low frequencies were identified as functionally important but were absent in the NFE/EUR and SAS populations, and no current records in literature support their role in TB drug PGx (Table 4, Supplementary Tables 4 and 5). Two variants in *AADAC* were identified as functionally important (Table 4). The 274C>T (rs760383318) SNP occurred only in the KHM population (0.0135) and could be of importance given a high CADD score (>27). Four variants in *CYP3A4* were predicted to be functionally important (Table 3 and 4), including the *CY3A4*11* (rs67784355), *CYP3A4*23* (rs57409622), *CYP3A4*7* (rs56324128) and *CYP3A4* (rs754968125) variants. In the *NR1I2* (*PXR*) gene, variants rs12721607, rs768816150, rs144833620, rs371067095 and rs1194240746 could have a functional effect (Table 4). Additional variation in less important TB PGx genes, including *CYP2C9, CYP2C19*, *CYP2B6, CUX2, Fam65B* and *CNTN5*, is shown in Supplementary Tables 4 and 5.

## Discussion

Despite extensive research into the role of various PGx genes in TB treatment outcomes and AT-DILI, the TB PGx landscape in African populations remains largely unexplored, particularly with respect to haplotype structures and variant frequencies in specific groups. To investigate TB PGx diversity enriched in African individuals and populations, we predicted functional genetic variation in ten TB PGx genes across 20 African populations using bioinformatic tools and determined star allele haplotype distributions for *NAT2.* We used SIFT, PolyPhen, Alpha Missense and CADD to distinguish functionally important variation; and used Project HOPE to verify predictions of selected variants; and PyPGx to identify haplotypes. Of the 34 prioritized variants, 30 were present exclusively or more frequently in more than one African population, compared with other global populations (Tables 3 and 4). Most variants had minor allele frequencies between 0.1 and 0.01 (Figure 2A). These frequency results are consistent with previous studies investigating ADME genes in African populations^28,67^. Similarly, the distribution of *NAT2* alleles and metabolizer phenotypes is unique to each African population investigated. Although the clinical utility of some of these variants remains uncertain, their low frequency and restriction to specific population groups does not preclude a non-negligible effect size and consequential effect on treatment outcomes. Indeed, rarity is often associated with greater functional impact and constitutes a large part of PGx variation. Focusing on the important TB PGx genes, we identify slow acetylator *NAT2* haplotypes and variants with predicted functional effects that could shape the PGx landscape of TB treatment in African populations in this study.

We included known TB PGx variation in our functional analysis to evaluate the predictive value and accuracy of our approach. As expected, VEP identified variants with established functional effects, and/or variants previously associated with TB PGx and ATDILI (Table 2). For *NAT2*, VEP correctly identified the SNPs defining the slow acetylator alleles *NAT2*14* (rs1801279) and *NAT2*6* (rs1799930), both of which are strongly associated with reduced INH metabolism^6,8,48^. Furthermore, the validated PGx TB variant rs4149056, which forms part of *SLCO1B1*5/*15 and *17*^68^ was also correctly classified as functionally relevant^34^. However, the SNPs defining the slow acetylator alleles *NAT2*5* (rs1801280) or *NAT2*7* (rs1799931) did not meet the downstream selection threshold (CADD PHRED>20), despite their well-established association with reduced INH metabolism^69^. Similarly, *NAT2*33*, *NAT2*39* and *NAT2*42* (Table 5) were not predicted to be functionally relevant by tools used in this study – although classified as poor, poor and indeterminate metabolizer alleles, respectively. None of the other variants listed in Table 2, met our criteria for functional importance. These results were not entirely unexpected, as in silico predictions routinely do not align with in situ findings^70^. This highlights that bioinformatic predictions alone cannot be considered definitive for assessing functional impact, that experimental validation is essential, and that perhaps, better tools are desirable to assess the more variation-tolerant ADME genes in particular^71^.

The *NAT2* gene encodes the enzyme responsible for approximately 88% of INH metabolism^72^, and thus plays a dominant role in TB PGx. The allele-defining SNPs *NAT2*31 (rs45477599), NAT2\**37 (rs *rs138707146*), and *NAT2*38* (rs55700793) are observed almost exclusively in African populations, albeit at low minor allele frequencies (up to 0.0176, 0.0096, and 0.018, respectively), and have been classified as poor metabolizing alleles. Alleles of indeterminate function such as *NAT2*43* and *NAT2*44* were also identified here, with frequencies of >1% in at least one population. The common alleles *NAT2*\**4,*5,*6 and *7* and *NAT2*14* currently provide a simple yet sufficient panel for predicting INH drug response^48,73^. However, evidence from the inclusion of the allele *NAT2*14*^74^ in the South African Zulu, suggests that incorporating additional variants such as known slow acetylator alleles *NAT2*31*, *NAT2*37*, *NAT2*38* and alleles of yet unidentified function could enhance predictive accuracy of acetylation phenotypes in some African populations^74^.

Some discrepancies between existing knowledge and the findings of our study need to be noted. Two allele-defining variants (*NAT2*50* (7A>G, rs200893121) and *NAT2*47* (683C>T, rs45518335)) which had previously been found in the YRI and LWK populations, respectively^28^, were not genotyped here in any of the African populations. We were unable to conclusively derive recently updated alleles *NAT2*30* (poor) *NAT2*34 (poor)*, *NAT2*36 (poor)*, *NAT2*40* (poor) and *NAT2*41* (indeterminate) with PyPGx, as these alleles are not part of the PyPGx database and share allele-defining SNPs (Table 5).

Although the use of PyPGx as a dated allele caller signifies a limitation, our study provides new data on the acetylation profiles for the Southern African NAM and KHM populations, which carry a substantially higher proportion of fast metabolizers than other African populations (Figure 3B). In the NAM, the rapid acetylator phenotype is seen in about 40% of the population, which is significantly higher than in the East African populations (Kenya KEMRI (KENK), Kenya Moi (KENM) and Uganda (UGA)) at < 10% (Figure 3B). This could bring about a higher risk of sub-therapeutic INH exposures, but equally, a lesser risk for AT-DILI in these South African populations compared to Eastern and Northern African population groups. Our results differ from a recent study by Malinga et al (2025)^28^, describing a relatively lower proportion of 10% rapid acetylators in a combined South African population group, consisting of mostly individuals of Bantu-speaking origin. Our study suggests the South African NAM and KHM have a unique acetylator profile, which may differ significantly from other populations in the same geographic region. The five-way admixed South African Coloured (SAC) population^75^, carries a smaller proportion of predicted rapid acetylators (>20%), but still a higher proportion than other African populations (Figure 3B)^14^. The NAT2*45 allele has never been reported in other African populations ^28^, but occurs in the SAC population (Table 5). At a very low frequency of 0.006%, it likely owes its presence in this unique population to European admixture^75^.

We identified potentially important African-specific variants in genes directly involved in TB drug metabolism and/or AT-DILI, including *SLCO1B1* (758G>A, 242T>C), *CYP2E1* (1370A>T, 646C>G), *GSTM1* (10480G>A), *GSTT1* (270759G>A), *GSTP1* (431T>C). Of all TB PGx genes, the *SLCO1B1* gene (in particular, *SLCO1B1*15*) exhibits the strongest association with RIF bioavailability^15–18^. Additional function-altering variants were observed in *AADAC*, *CYP3A4*, *CES2* and other genes (Table 4, Supplementary Table 4 and 5) which are also implicated in AT-DILI. Although the very low frequencies of these variants make their large-scale contribution uncertain, our findings underscore that current knowledge of TB PGx does not adequately capture the PGx variation in African populations. Because some populations were represented only by imputed genotyped data and certain groups in our study had small sample sizes (with just 13 individuals for the EGY), additional, uncharacterized variation was likely not captured (recorded as “NA” in Figure 2A), indicating that our study may underestimate the true diversity within these groups. The population diversity, rarity, and uncertain functional impact of PGx variants underscore the need for broadly inclusive genotyping panels in African populations. Alternatively, NGS presents a more holistic approach towards conducting individualized drug therapy^9^. The single coding exon 2 of the *NAT2* gene, for example, would be a suitable candidate for sequence analysis, stretching over only 870 bp.

Some variation enriched in African populations might have been missed by implementing the widely accepted CADD threshold >20. We did not include copy number variants (CNVs), despite their role in determining alleles particularly in *GST* enzymes. To date, the ability of computational algorithms to detect and assess the effect of intronic variants and CNV effects computationally has been limited, especially in ADME genes. However, the larger the effect size of a variant, the more likely it is detectable by in-silico functional assessment. Notably, the variants that were correctly identified demonstrate a large effect size in TB PGx (for example, *NAT2*14*).

In addition to changes in protein structure and activity, genes are altered at the transcriptional level. The PXR nuclear receptor gene regulates the transcription of other important PGx genes (e.g. *CYP3A4, SLCO1B1 and AADAC*) in response to ligands such as RIF. Variants in *PXR* are thus expected to only play an indirect role in regulating TB drug metabolism. The small effect size (and low frequency) of these variants makes them less important specifically for large-scale TB PGx. However, they could influence general drug metabolism. While we addressed knowledge gaps in TB PGx in African populations, the genes investigated are also central to other drug therapies. For instance, actionable variants in *SLCO1B1* influence atorvastatin and simvastatin bioavailability, accounting for ∼30% of drug prescriptions and ∼10% of drug expenditure in South Africa^26^. Our study offers novel insights into the distribution of *NAT2* star alleles in the Southern African NAM, KHM, SAC and XHO populations, some of which had not previously been documented^76,77^.

An estimated ∼95% of all individuals across ancestries carry ≥1 PGx variant, and ∼50% ≥2 actionable variants^78^. The comparatively higher density of variation in ADME genes observed in African populations^28^ supports the assumption that the number of actionable variants will be even greater in African groups than in non-African populations, even though variants are often rare, population-specific and in some cases not yet validated. The PREPARE study demonstrated that pre-emptive, genotype-guided dosing and drug selection can reduce drug-induced ADRs by 30%^79^. Similar studies in South Africa, focusing on commonly prescribed actionable drugs, suggest that integrating genomic medicine into African healthcare systems could lower ADR incidence to a similar extent and ease financial pressure on healthcare systems^26,80^. TB drugs are not currently classified actionable and were therefore excluded from these studies, despite their substantial prescription rates. The high burden of TB/HIV co-infection in Africa, where patients often receive chronic multi-drug therapy for several months, together with rising drug resistance and the 18-30% of TB patients experiencing AT-DILI^81^, underscores the need to elucidate the variability underlying TB PGx.

Several initiatives are laying the groundwork for cataloguing the diversity of the general PGx landscape in Africa^22,28,82–84^. Building on this progress, further efforts to enhance data sharing, reporting and safeguarding - along with healthcare provider training and guideline development – will strengthen the foundation for targeted pilot studies. With these advances, governments, patients and healthcare systems across Africa are well positioned to benefit from optimized treatments, particularly for TB. By embracing cultural, environmental, and genetic diversity, inclusive, precise and effective care can be achieved.

## Data Availability

All data produced in the present study are available upon reasonable request to the authors
All data produced in the present work are contained in the manuscript

## List of abbreviations

AADAC: arylacetamide deacetylase
ADME: absorption, distribution, metabolism and excretion
ADR: adverse drug reaction
AGR: African Genome Resource
AT-DILI: anti-TB drug-induced liver injury
BDQ: bedaquiline
BFA: Burkina Faso
CADD: combined annotation dependent depletion
CAM: Cameroon
CES2: carboxyl-esterase 2
CPIC: clinical pharmacogenetics implementation consortium
CYP2E1: cytochrome P450 family 2 subfamily E member 1
CYP3A4: cytochrome P450 family 3 subfamily A member 4
EGY: Egypt
ETH: Ethiopia
EUR/NFE: European and Non-Finnish
GHA: Ghana
GSTM1: glutathione S-transferase Mu 1
GSTP1: glutathione S-transferase Pi 1
GSTT1: glutathione S-transferase Theta 1
INH: isoniazid
KENK: Kenya KEMRI
KENM: Kenya Moi
KHM: Khomani
LWK: Luhya Western Kenya
MAF: minor allele frequency
MAF: minor allele frequency
MAL: Malawi
MSL: Mende Sierra Leone
MZB: Mozabite
NAM: Nama
NAT2: N-acetyl transferase 2
PCR: polymerase chain reaction
PGx: Pharmacogenetic
PolyPhen: Polymorphism Phenotyping
PXR: Pregnane X Receptor
PYG: Pygmy
PZA: pyrazinamide
QC: quality control
RIF: rifampicin
SAC: South African Coloured
SAS: South Asian
SIFT: Sorting Intolerant From Tolerant
SLCO1B1: solute carrier organic anion transporter family member 1B1
SNP: single nucleotide polymorphism
TAN: Tanzania
TB: tuberculosis
UGA: Uganda
VCF: variant call format
VEP: Variant Effect Predictor
XHO: Xhosa
YRI: Yoruba Nigeria

