## Supplementary Tables 1-5 for "In-silico functional prediction of novel tuberculosis pharmacogenetic variants and NAT2 phenotype prediction in African populations"

*Supplementary Table 1: PyPGx-predicted NAT2 allele frequencies across African populations, indicating the number of individuals in each population in brackets. For the populations indicated in grey, no data was available for the NAT2*5 allele-defining SNP. The missing NAT2*5 allele prohibited prediction of phenotypes for the YRI, ESN, LWK, TAN and MSL population, indicated in grey. Insufficient data and small sample numbers led to exclusion of populations MZB, GHA and ETH.*

| | BFA (122) | CAM (131) | PYG (125) | MAL (115) | UGA (85) | KENK (187) | KENM (173) | XHO (164) | KHM (185) | NAM (127) | SAC (82) |
|---|---|---|---|---|---|---|---|---|---|---|---|
| *1 (legacy *4) (rapid) | 0.0 | 0.0 | 6.4 | 0.0 | 0.0 | 0.0 | 8.7 | 8.8 | 19.5 | 26.8 | 0.0 |
| ***5 (slow)** | 31.6 | 31.3 | 24.8 | 32.2 | 35.3 | 35.3 | 42.5 | 28.7 | 19.5 | 18.9 | 29.2 |
| ***6 (slow)** | 24.2 | 24.1 | 22.0 | 24.8 | 27.7 | 26.7 | 28.0 | 23.3 | 14.3 | 16.5 | 23.4 |
| ***7 (slow)** | 3.3 | 1.5 | 1.6 | 2.2 | 2.4 | 2.9 | 2.6 | 1.0 | 3.5 | 1.2 | 4.8 |
| *1 (legacy *12) (rapid) | 16.0 | 21.4 | 27.2 | 17.0 | 15.3 | 15.8 | 7.8 | 17.9 | 37.0 | 32.3 | 38.7 |
| *4 (legacy *13) (rapid) | 14.3 | 10.3 | 9.2 | 9.1 | 7.7 | 8.3 | 4.9 | 13.9 | 3.5 | 2.4 | 0.0 |
| *14 (slow) | 9.8 | 9.5 | 8.0 | 12.2 | 10.0 | 9.6 | 4.3 | 6.4 | 2.2 | 2.0 | 3.8 |
| *43 (legacy *22) (intdet.) | 0.8 | 0.8 | 0.8 | 1.3 | 0.0 | 0.5 | 0.0 | 0.0 | 0.0 | 0.0 | 0.3 |
| *31 (legacy *23) (slow) | 0.0 | 0.0 | 0.0 | 0.0 | 0.6 | 0.3 | 0.3 | 0.0 | 0.0 | 0.0 | 0.0 |
| *44 (legacy *24) (indet.) | 0.0 | 1.2 | 0.0 | 1.3 | 1.2 | 0.5 | 0.9 | 0.0 | 0.5 | 0.0 | 0.0 |

*Supplementary Table 2: PyPGx-predicted NAT2 metabolizer phenotypes across African population groups, indicating the number of each individual in each population in brackets. The missing NAT2*5 allele prohibited prediction of phenotypes for the YRI, ESN, LWK, TAN and MSL populations. Insufficient data and small sample numbers led to exclusion of populations MZB, GHA and ETH.*

| Phenotype | BFA (122) | CAM (131) | PYG (125) | MAL (115) | UGA (85) | KENK (187) | KENM (173) | XHO (164) | KHM (185) | NAM (127) | SAC (82) |
|---|---|---|---|---|---|---|---|---|---|---|---|
| **rapid** | 0.07 | 0.12 | 0.20 | 0.09 | 0.05 | 0.05 | 0.05 | 0.13 | 0.36 | 0.43 | 0.17 |
| **intermediate** | 0.45 | 0.36 | 0.44 | 0.33 | 0.35 | 0.36 | 0.34 | 0.55 | 0.47 | 0.39 | 0.44 |
| **poor** | 0.46 | 0.48 | 0.34 | 0.51 | 0.58 | 0.56 | 0.60 | 0.32 | 0.16 | 0.19 | 0.39 |
| **indeterminate** | 0.02 | 0.04 | 0.02 | 0.07 | 0.02 | 0.02 | 0.02 | 0.00 | 0.01 | 0.00 | 0.01 |

*Supplementary Table 3: Functional assessment of point mutations on protein structure in Project HOPE.*

| Gene / UniProt | Nucleotide / Amino Acid / RS ID | Project HOPE |
|---|---|---|
| *NAT2*31* (P11245) | 70 T>A / Leu 24 Ile / rs45477599 | The mutation residue does not prefer α-helices as secondary structure. However, residues that have some properties in common with your mutated residue were observed. This means that in some rare cases your mutation might occur without damaging the protein. HOPE cannot find a structural effect for this mutation. |
| *NAT2*38* (P11245) | 766 A>G / Lys 256 Glu / rs55700793 | stop codon |
| *NAT2*37* (P11245) | 638 C>T / Pro 213 Leu / rs138707146 | The residue is in a binding site, and the mutant amino acid is larger than the wild-type, potentially disturbing protein stability. Wild-type residue is very conserved, mutant likely damaging. |
| *SLCO1B1* (Q9Y6L6) | 758 G>A / Arg 253 Gln / rs11045853 | The wild-type residue charge was POSITIVE, the mutant residue charge is NEUTRAL, probably not damaging to the protein, but can cause loss of interactions with other molecules or residues. The mutant residue is smaller; this might lead to loss of interactions. |
| *SLCO1B1* (Q9Y6L6) | 242 T>C / Ile 81 Thr / rs1353535924 | The mutant residue is smaller and less hydrophobic. MetaRNN score is 0.9466, indicating high pathogenicity. Residues that have some properties in common with the wild type, were observed. |
| *CYP2E1* (P05181) | 517 G>T / Gly 173 Ser / rs60452492 | The mutant residue is bigger than the wild-type residue and will not fit the core of the protein. Wild-type glycine is flexible to generate torsion angles; mutation will force the local backbone into an incorrect conformation and will disturb the local structure. |
| *CYP2E1* (P05181) | 1370 A>T / His 457 Arg / rs28969387 | Based on the conservation information the mutation is probably not damaging to the protein. The mutated residue is located in a domain that is important for the activity of the protein and in contact with residues in another domain. It is possible that this interaction is important for the correct function of the protein. The mutant residue introduces a charge in a buried residue which can lead to protein folding problems. |
| *CYP2E1* (P05181) | 646C>G / Gln216Glu / rs1194608147 | Wild-type residue is neutral, mutant is negative, could disturb interaction between two domains. |
| *CYP2C19* (P33261) | 55 C>T / Ala 350 Thr / rs201509150 | The mutated residue is in a domain important for binding molecules. The mutated residue is in contact with residues in another domain, possibly disturbing these contacts. The bigger mutant probably will not fit into the core, cause loss of hydrophobic interactions and does not prefer α-helices as secondary structure. Neither your mutant residue nor another residue type with similar properties was observed at this position in other homologous sequences. Based on conservation scores this mutation is probably damaging to the protein. |

| | | |
|---|---|---|
| *GSTT1* (P30711) | 562 C>T / Arg 188 Ter / rs144686326 | stop codon |
| *GSTP1* (P09211) | 431T>C / Ile144Thr / (rs199949339) | Mutant residue is smaller and might cause loss of hydrophobic interactions within the protein. Other residues have been observed at this site; mutant may likely be benign. |
| *GSTM1* (P09488) | 10480G>A/ Ala213Thr / rs533860247 | Mutant residue is bigger, more hydrophobic than the wild-type, which is in an α-helix. The mutant does not prefer this secondary structure. This site is 100% conserved, indicating that the mutant residue is likely pathogenic. |
| *CYP2C9* (P11712) | 538T>C / Ser 180 Pro / rs367922573 | Mutant residue is bigger and more hydrophobic than the wild type and disrupts a hydrogen bond and α-helix structure. No similar residues were observed in this position; mutant is likely damaging. |

*Supplementary Table 4: Variation in additional TB PGx genes with a predicted functional effect, with PubMed records. Populations were excluded if data was missing.*

| | **SIFT** | **PolyPhen** | **CADD PHRED** | **Alpha Missense** | **Clinical significance** | **Frequencies across African populations** | Frequencies across Non-African populations | African populations with frequencies > 0.01 |
|---|---|---|---|---|---|---|---|---|
| *CYP2C9*2,* 430C>T, rs1799853, | **0.05** | **0.986** | **21.2** | **0.2301** | **drug response** | 0 (BFA), 0.0769 (EGY), 0.2447 (ETH), 0 (GHA), 0.0048 (GWD), 0.008 (KENK), 0.0058 (KENM), 0.0081 (KHM), 0 (LWK), 0.0058 (KENM), 0.0087 (MAL), 0 (MSL), 0.1667 (MZB), 0.006 (NAM), 0 (PYG), 0.027 (SAC), 0.1694 (TAN), 0 (UGA), 0 (YRI), 0.003 (XHO) | 0.124 (EUR/NFE), 0.035 (SAS) | 5 |
| *CYP2C9*9,* 752A>G, rs2256871 | **0** | 0.821 | **20.9** | **0.2429** | benign | 0 (EGY), 0.0319 (ETH), 0.0336 (GWD), 0.1123 (KENK), 0.1127 (KENM), 0.1 (KHM), 0.1524 (LWK), 0.0267 (MSL), 0.0185 (MZB), 0.1667 (NAM), 0.108 (PYG), 0.1882 (UGA), 0.0926 (YRI), 0.1341 (XHO) | 0.001 (EUR/NFE), 0 (SAS) | 13 |
| *CYP2C9*31,* 980T>C, rs57505750 | **0** | 1 | **23.3** | **0.3094** | uncertain | 0 (CAM), 0 (EGY), 0 (GHA), 0 (GWD), 0.0173 (KENM), 0 (KHM), 0.0122 (LWK), 0 (MAL), 0 (MSL), 0 (PYG), 0.0006 (SAC), 0 (YRI), 0.0152 (XHO) | 0 (EUR/NFE), 0 (SAS) | 3 |
| *CYP2C9*11,* 1003C>T, rs28371685 | **0** | **0.998** | **22.2** | **0.2408** | **drug response** likely benign | 0.0574 (BFA), 0.0115 (CAM), 0 (EGY), 0.0354 (GHA), 0.0385 (GWD), 0.0267 (KENK), 0.026 (KENM), 0.0027 (KHM), 0.0183 (LWK), 0.0348 (MAL), 0.02 (MSL), 0.0185 (MZB), 0.092 (PYG), 0.0043 (SAC), 0.0289 (TAN), 0.0118 (UGA), 0.0509 (YRI), 0.0091 (XHO) | 0.0020 (EUR/NFE), 0.001 (SAS) | 14 |

| | | | | | | | | |
|---|---|---|---|---|---|---|---|---|
| *CYP2C19*9,* 431G>C, rs17884712 | **0.01** | **0.982** | **22** | **0.1736** | **drug response** | 0 (EGY), 0.0106 (ETH), 0.0096 (GWD), 0.0107 (KENK), 0.0087 (KENM), 0.1378 (KHM), 0.0061 (LWK), 0.0087 (MAL), 0.02 (MSL), 0 (MZB), 0.1726 (NAM), 0.024 (PYG), 0.0607 (SAC), 0.0059 (UGA), 0.0046 (YRI), 0.0671 (XHO) | 0 (EUR/NFE), 0.0001 (SAS) | 8 |
| *CYP2C19,* 556C>T, rs183701923 | **0.02** | **0.855** | **23** | **0.2663** | **no protein expression** | 0 (EGY), 0 (GHA), 0 (GWD), 0 (KENK), 0 (KENM), 0.0061 (LWK), 0.0087 (MAL), 0 (MSL), 0 (SAC), 0 (UGA), 0 (YRI), 0 (XHO) | 0 (EUR/NFE), 0 (SAS) | 0 |
| *CYP2C19*10,* 680C>T, rs6413438 | **0** | **0.995** | **21.5** | **0.2916** | **drug response** | 0.0123 (BFA), 0 (CAM), 0 (EGY), 0 (ETH), 0.0044 (GHA), 0.0048 (GWD), 0 (KENM), 0 (KHM), 0 (LWK), 0 (MSL), 0 (PYG), 0 (YRI), 0 (XHO) | 1 (EUR/NFE), 0 (SAS) | 1 |
| *CYP2C19,* 1048G>A, rs201509150 | **0.01** | **0.97** | **25.5** | 0.7374 | uncertain | 0.0185 (MZB) | 0.0001 (EUR/NFE), 0 (SAS) | 1 |
| *CYP2C19*45,* 394C>T, rs149590953 | **0.02** | 0.814 | **20.1** | NA | NA | 0.003 (XHO) | 0.0001 (EUR/NFE), 0.0004 (SAS) | 0 |
| *CYP2C19,* 784G>A, rs577255883 | **0.01** | 0.603 | **22.6** | 0.6524 | NA | 0.0012 (SAC) | 0.0001 (EUR/NFE), 0.0068 (SAS) | 0 |

*Supplementary Table 5: Variation in additional TB PGx genes with a predicted functional effect, without PubMed records. Populations were excluded if data was missing.*

| | **SIFT** | **PolyPhen** | **CADD PHRED** | **Frequencies across African populations** | **African populations with frequencies > 0.01** |
|---|---|---|---|---|---|
| *CYP2B6,* rs757472041, 19:41510240 | 0 | 0.874 | 24 | 0 (ETH), 0 (KENK), 0 (KENM), 0 (PYG), 0.006 (UGA), 0 (KHM), 0 (XHO) | 0 |
| *CYP2B6,* rs760463009, 19:41515953 | 0.23 | 0.990 | 20 | 0 (BFA), 0.003 (KENM), 0 (KHM), 0 (MAL), 0 (PYG), 0 (TAN), 0 (XHO) | 0 |
| *CUX2,* rs576939929, 12:111472033 | 0.04 | 0.398 | 25.5 | 0.005 (GWD), 0 (LWK), 0 (YRI), 0 (MSL), 0 (SAC) | 0 |
| *CUX2,* rs201396910, 12:111652085 | 0.01 | 0.732 | 32 | 0 (BFA), 0.005 (GWD), 0 (LWK), 0 (PYG), 0 (YRI), 0.007 (MSL), 0 (SAC), 0 (XHO) | 0 |
| *CUX2,* rs868561431, 12:111701615 | 0 | 0.617 | 22.5 | 0.011 (ETH), 0 (KENK), 0 (KENM), 0 (KHM), 0 (MAL), 0 (PYG), 0 (TAN), 0 (UGA), 0 (XHO) | 1 |
| *CUX2,* rs202242120, 12:111731300 | 0.03 | 0 | 22.2 | 0.004 (BFA), 0 (ETH), 0 (GWD), 0.011 (KENK), 0.009 (KENM), 0.005 (KHM), 0.012 (LWK), 0.009 (MAL), 0.012 (PYG), 0.017 (TAN), 0.006 (UGA), 0.009 (YRI), 0.027 (MSL), 0.007 (SAC), 0.003 (XHO) | 4 |

| | | | | | |
|---|---|---|---|---|---|
| *CUX2,* rs562432690, 12:111758410 | 0 | 0.007 | 24 | 0 (BFA), 0 (ETH), 0.005 (GWD), 0 (KENK), 0.003 (KENM), 0 (KHM), 0 (LWK), 0 (MAL), 0 (PYG), 0 (TAN), 0 (UGA), 0 (YRI), 0 (MSL), 0.002 (SAC), 0 (XHO) | 0 |
| *CUX2,* rs568862964, 12:111785423 | 0.03 | 0.001 | 21.3 | 0.004 (BFA), 0 (ETH), 0 (KENM), 0 (MAL), 0 (SAC), 0 (XHO) | 0 |
| *FAM65B,* rs373913240, 6:24806612 | 0 | 0.994 | 24 | 0.012 (BFA), 0.015 (CAM), 0 (ETH), 0.004 (GHA), 0.005 (GWD), 0.008 (KENK), 0.003 (KENM), 0.006 (LWK), 0.009 (MAL), 0 (MZB), 0.008 (PYG), 0 (UGA), 0 (YRI), 0.027 (MSL) | 3 |
| *FAM65B,* rs930774934, 6:24809966 | 0.36 | 0.987 | 22.3 | 0 (BFA), 0 (CAM), 0 (ETH), 0 (GHA), 0.008 (KENK), 0.003 (KENM), 0 (KHM), 0.004 (MAL), 0.004 (PYG), 0 (UGA), 0.021 (XHO) | 1 |
| *FAM65B,* rs181654531, 6:24825482 | 0.01 | 0.572 | 26.3 | 0 (ETH), 0 (GWD), 0 (KENM), 0 (KHM), 0 (LWK), 0 (PYG), 0.005 (YRI), 0.007 (MSL), 0 (SAC), 0 (XHO) | 0 |
| *FAM65B,* rs115013548, 6:24843097 | 0.06 | 0.636 | 24.3 | 0.025 (BFA), 0.015 (CAM), 0.011 (ETH), 0.009 (GHA), 0.024 (GWD), 0 (KENK), 0.006 (KENM), 0.014 (KHM), 0 (LWK), 0 (MZB), 0.035 (UGA), 0.028 (YRI), 0.027 (MSL), 0.024 (SAC), 0.006 (XHO) | 9 |
| *FAM65B*, rs11967003, 6:24865659 | 0.19 | 0.998 | 23.7 | 0.119 (BFA), 0.103 (CAM), 0.021 (ETH), 0.106 (GHA), 0.005 (GWD), 0.067 (KENK), 0.064 (KENM), 0.038 (KHM), 0.049 (LWK), 0.087 (MAL), 0 (MZB), 0.053 (UGA), 0.083 (YRI), 0.12 (MSL), 0.06 (NAM), 0.085 (XHO) | 14 |
| *FAM65B,* rs771443041, 6:24874015 | 0.01 | 0.994 | 24.4 | 0 (BFA), 0 (GHA), 0 (KENK), 0.003 (KENM), 0 (MAL), 0 (UGA), 0 (XHO) | 0 |
| *FAM65B,* rs530256583, 6:24874017 | 0.06 | 0.998 | 25.4 | 0 (BFA), 0 (GHA), 0 (GWD), 0 (KENK), 0.003 (KENM), 0 (LWK), 0 (MAL), 0 (UGA), 0 (YRI), 0 (MSL), 0 (SAC), 0 (XHO) | 0 |
| *CNTN5,* rs137976542, 11:99715682 | 0.04 | 0.830 | 25.4 | 0 (CAM), 0 (ETH), 0.004 (GHA), 0.01 (GWD), 0 (KENK), 0.012 (KENM), 0.024 (LWK), 0 (MAL), 0 (MZB), 0.004 (PYG), 0.012 (UGA), 0 (YRI), 0.007 (MSL), 0 (XHO) | 4 |
| *CNTN5,* rs201910584, 11:99931945 | 0 | 0.995 | 29 | 0.093 (MZB) | 1 |
| *CNTN5,* rs200782641, 11:99944948 | 0 | 0.410 | 24.4 | 0 (BFA), 0.004 (CAM), 0 (ETH), 0 (GWD), 0 (KENK), 0 (KENM), 0 (KHM), 0 (LWK), 0 (MAL), 0 (PYG), 0 (UGA), 0 (YRI), 0 (MSL), 0 (SAC), 0 (XHO) | 0 |
| *CNTN5*, rs538146525, 11:100064349 | 0.36 | 0.984 | 25.2 | 0.003 (KENK), 0 (KENM), 0.003 (KHM), 0 (PYG), 0 (UGA), 0 (SAC), 0 (XHO) | 0 |
| *CNTN5,* rs200512456, 11:100169923 | 0.12 | 0.994 | 23.1 | 0.011 (ETH), 0 (GWD), 0 (KENK), 0 (KENM), 0.008 (KHM), 0 (LWK), 0 (MZB), 0 (UGA), 0 (YRI), 0 (MSL), 0.001 (SAC), 0 (XHO) | 1 |
| *CNTN5,* rs200134748, 11:100211896 | 0.01 | 0.995 | 25.8 | 0 (BFA), 0(ETH), 0.014 (GWD), 0 (KENK), 0 (KENM), 0 (KHM), 0 (LWK), 0 (MZB), 0 (PYG), 0(UGA), 0(YRI), 0(MSL), 0(SAC), 0(XHO) | 1 |
| *CNTN5,* rs369992087, 11:100211909 | 0.02 | 0.448 | 23.9 | 0 (ETH), 0 (KENK), 0.003 (KENM), 0 (KHM), 0.004 (MAL), 0 (PYG), 0 (UGA), 0 (XHO) | 0 |
